# COVID-19 Vaccine Acceptance and Hesitancy in Low and Middle Income Countries, and Implications for Messaging

**DOI:** 10.1101/2021.03.11.21253419

**Authors:** Julio S. Solís Arce, Shana S. Warren, Niccoló F. Meriggi, Alexandra Scacco, Nina McMurry, Maarten Voors, Georgiy Syunyaev, Amyn Abdul Malik, Samya Aboutajdine, Alex Armand, Saher Asad, Britta Augsburg, Antonella Bancalari, Martina Björkman Nyqvist, Ekaterina Borisova, Constantin Manuel Bosancianu, Ali Cheema, Elliott Collins, Ahsan Zia Farooqi, Mattia Fracchia, Andrea Guariso, Ali Hasanain, Anthony Kamwesigye, Sarah Kreps, Madison Levine, Rebecca Littman, Melina Platas, Vasudha Ramakrishna, Jacob N. Shapiro, Jakob Svensson, Corey Vernot, Pedro C. Vicente, Laurin B Weissinger, Baobao Zhang, Dean Karlan, Michael Callen, Matthieu Teachout, Macartan Humphreys, Saad B. Omer, Ahmed Mushfiq Mobarak

**Author notes:** Corresponding author. 1 Church Street, New Haven, CT 06510. Corresponding author. 165 Whitney Avenue, New Haven, CT 06520-8200. First author (Julio S. Solís Arce, Shana S. Warren, Niccoló F. Meriggi, Alexandra Scacco, Nina McMurry, Maarten Voors, Georgiy Syunyaev, Amyn Abdul Malik). Last author (Dean Karlan, Michael Callen, Matthieu Teachout, Macartan Humphreys).

## Abstract

We analyze COVID-19 vaccine acceptance across 15 survey samples covering ten low- and middle-income countries (LMICs) in Asia, Africa, and South America, Russia (an upper-middle-income country), and the United States, using survey responses from 44,260 individuals. We find considerably higher willingness to take a COVID-19 vaccine in LMIC samples (80% on average) compared to the United States (65%) and Russia (30%). Vaccine acceptance was primarily explained by an interest in personal protection against COVID-19, while concern about side effects was the most commonly expressed reason for reluctance. Health workers were the most trusted sources of information about COVID-19 vaccines. Our findings suggest that prioritizing vaccine distribution to LMICs should yield high returns in promoting global immunization coverage, and that vaccination campaigns in these countries should focus on translating acceptance into uptake. Messaging highlighting vaccine efficacy and safety, delivered by healthcare workers, may be most effective in addressing remaining hesitancy.

---

A safe and effective vaccine against COVID-19 is a critical tool to control the pandemic. As of April 27, 2021, 22 vaccines had advanced to Stage 3 clinical trials^1^ and more than a dozen vaccines had been approved in multiple countries^2^. For example, as of April 27, 2021, Comirnaty (BioNTech/Pfizer) had been approved in more than 80 countries, while the AstraZeneca vaccine had the most country authorizations at 92.^2^ At present, however, global vaccine distribution remains highly unequal, with much of the current supply directed toward high-income countries.^3^

While effective and equitable distribution of COVID-19 vaccines is a key policy priority, ensuring the population’s acceptance is equally important. Trust in vaccines and the institutions that administer them are key determinants of the success of any vaccination campaign.^4^ Several studies have investigated willingness to take a potential COVID-19 vaccine in high-income countries,^5–10^ and some studies have included middle-income countries.^3, 11^ Less is known, however, about vaccine acceptance in low-income countries where large-scale vaccination has yet to begin. Understanding the drivers of COVID-19 vaccine acceptance is of global concern, since a lag in vaccination in any country may result in the emergence and spread of new variants that can overcome vaccine- and prior disease-conferred immunity.^12, 13^

Our study complements the emerging global picture of COVID-19 vaccine acceptance by focusing primarily on lower income countries. We construct a sample of low- and middle-income countries (LMICs) with wide geographic coverage across Africa, Asia and Latin America. We move beyond documenting vaccine acceptance rates and analyze data on the reasons for acceptance and hesitancy, which is critical for informing the design of effective vaccine distribution and messaging.

Acceptance of childhood vaccination for common diseases—such as measles (MCV), Bacille Calmette-Guérin (BCG) and diphtheria, tetanus, and pertussis (DTP)—is generally high in LMICs, providing cause for optimism about the prospects for COVID-19 vaccine uptake. Table 1 summarizes general vaccine acceptance and coverage rates of childhood vaccines in 2018, prior to the current pandemic, for the countries included in our study. Agreement on the importance of childhood vaccinations is markedly higher in these LMICs compared to Russia and the United States. Still, existing studies on COVID-19 vaccine acceptance document substantial variation, both across and within countries, including in settings with high background levels of other vaccinations.^3, 4, 11^

**Table 1:** Vaccination beliefs and coverage for the countries in our sample.

|  | % Respondents agreeing Vaccines are... |  |  | Vaccine coverage in 2019 (% of infants) |  |  | % of parents with any child that was ever vaccinated |
| --- | --- | --- | --- | --- | --- | --- | --- |
|  | Effective | Safe | Important for children to have | Tuberculosis (BCG) | Diphtheria, Tetanus and Pertussis (DTP1) | Measles (MCV1) |  |
| Burkina Faso | 87 | 72 | 95 | 98 | 95 | 88 | 97 |
| Colombia | 83 | 84 | 99 | 89 | 92 | 95 | 95 |
| India | 96 | 97 | 98 | 92 | 94 | 95 | 92 |
| Mozambique | 87 | 93 | 98 | 94 | 93 | 87 | 95 |
| Nepal | 89 | 93 | 99 | 96 | 96 | 92 | 95 |
| Nigeria | 82 | 92 | 96 | 67 | 65 | 54 | 95 |
| Pakistan | 91 | 92 | 95 | 88 | 86 | 75 | 94 |
| Rwanda | 99 | 97 | 99 | 98 | 99 | 96 | 100 |
| Sierra Leone | 95 | 95 | 99 | 86 | 95 | 93 | 97 |
| Uganda | 82 | 87 | 98 | 88 | 99 | 87 | 98 |
| Russia | 67 | 48 | 80 | 96 | 97 | 98 | 96 |
| USA | 85 | 73 | 87 | . | 97 | 90 | 95 |

Table 1 presents an overview of vaccination beliefs and incidence across countries in our sample. Columns 2-4 and 8 use data from the Wellcome Global Monitor 2018. Column 8 shows the percentage of respondents who are parents and report having had any of their children ever vaccinated. Columns 2-4 show the percentage of all respondents that either strongly agree or somewhat agree with the statement above each column. All percentages are obtained using national weights. Columns 5-7 use data from the World Health Organization on vaccine incidence. Columns 5-7 report the percentage of infants per country receiving the vaccine indicated in each column.

This literature cites concerns about COVID-19 vaccine safety, including concerns about the rapid pace of vaccine development, as reasons for hesitancy in higher-income settings,^3, 5^ but other reasons may feature more prominently in LMICs. For instance, reported COVID-19 mortality rates have been consistently lower in LMICs relative to higher income countries.^14^ If individuals feel the risk of disease is less serious, they may be less willing to accept any perceived risks of vaccination.^15^ Previous studies of healthcare utilization in LMICs have also highlighted factors such as healthcare quality concerns,^16^ negative historical experiences involving foreign actors,^17, 18^ weak support from traditional leaders,^19^ and mistrust in government^20^ as barriers to uptake, which could apply to COVID-19 vaccination as well.

To promote vaccination against COVID-19 and develop effective messaging strategies, we need to know whether people are willing to take COVID-19 vaccines, the reasons why they are willing or unwilling to do so, and the most trusted sources in their decision-making. Our study investigates these questions using a common set of survey items deployed across 13 studies in Africa, South Asia, and Latin America: seven in low-income countries (Burkina Faso, Mozambique, Rwanda, Sierra Leone, Uganda), five in lower-middle-income countries (India, Nepal, Nigeria, Pakistan), and one in an upper-middle-income country (Colombia). We compare these findings to those from two countries at the forefront of vaccine research and development, Russia and the United States. To select studies to include in our sample, we conducted an internal search within Innovations for Poverty Action (IPA), the International Growth Center (IGC), and the Berlin Social Science Center (WZB) for projects with plans to collect survey data in the second half of 2020. Study PIs agreed to include a set of common questions about COVID-19 vaccine attitudes. This strategy was guided by the need to collect information quickly and cost effectively using a survey modality (phone) that was both safe given pandemic conditions and appropriate for contexts with limited internet coverage. The final set of samples included in our study therefore reflects populations that fall under the current research priorities at IGC, IPA and WZB and, in the case of IPA and IGC, donors that prioritize working in the Global South.

## Results

Our main results are shown in Figure 1 and reproduced as Appendix A Table 4. The first column provides overall acceptance rates in each study, while the remaining columns disaggregate acceptance rates by respondent characteristics. The “All LMICs” row reports averages for LMIC countries and excludes Russia and the USA.

**Figure 1:**
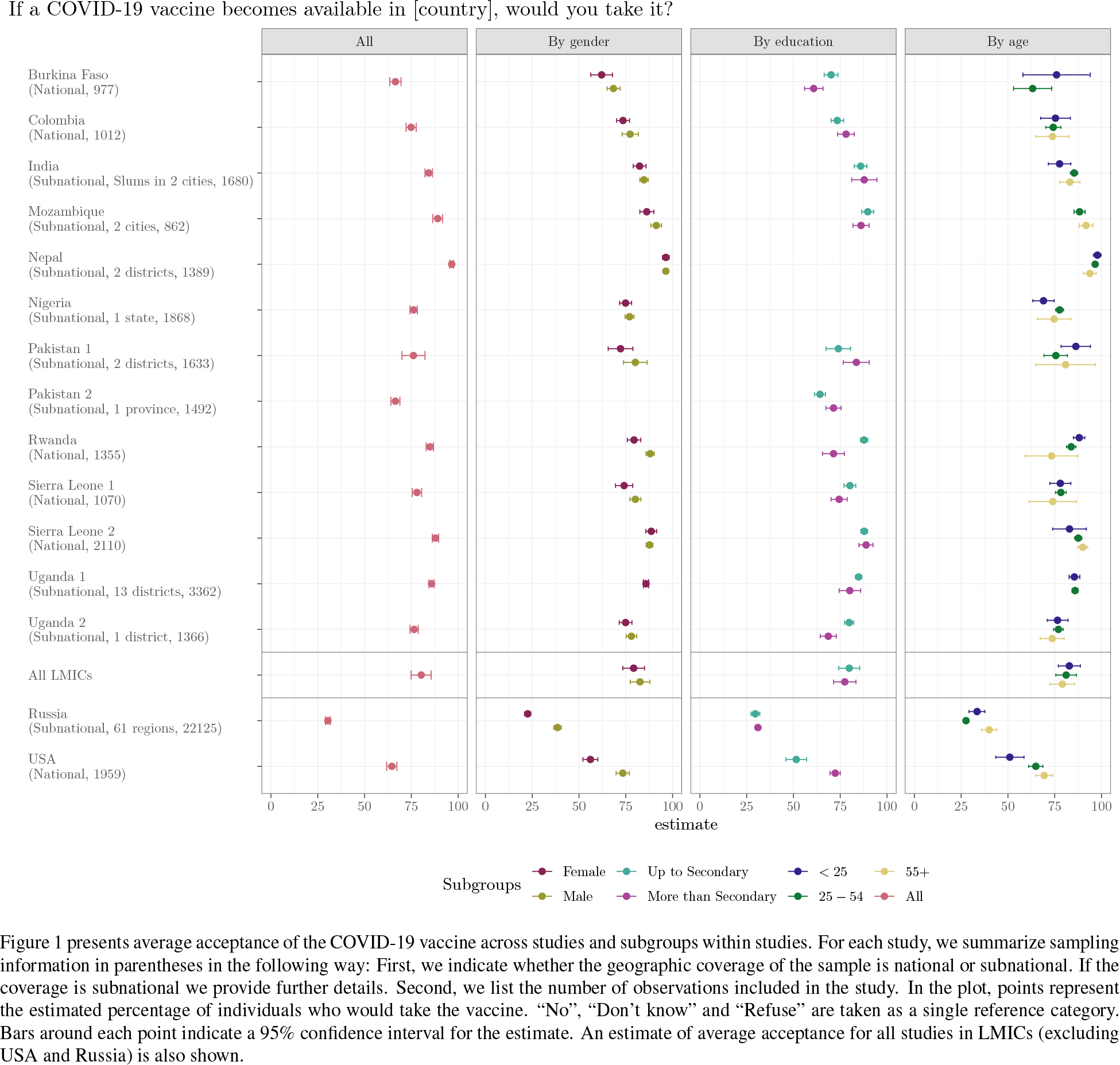
Acceptance rates overall and broken down by respondent characteristics.

We document meaningful variation in vaccine acceptance across and within LMICs, but generally high levels of acceptance in LMICs overall. The average acceptance rate across studies was 80.3% (95% CI 74.9–85.6%), with a median of 78, a range of 30.1 percent points and an interquartile range of 9.7. Our estimate of *τ*^2^ is 0.007 which implies a standard deviation over country averages of 0.084.

The acceptance rate in every LMIC sample was higher than in the USA (64.6%, 61.8–67.3%) and Russia (30.4%, 29.1–31.7%). Reported acceptance was lowest in Burkina Faso (66.5% 63.5– 69.5%) and Pakistan 2 (66.5%, 64.1–68.9%). The case of Pakistan may be linked to negative historical experiences with foreign-led vaccination campaigns.^21, 22^ This hesitancy may be particularly problematic, given the magnitude of the second wave in neighboring India just as vaccines are becoming available, with accelerating cases in Bangladesh, Pakistan and India threatening to overwhelm health infrastructure.

We find limited evidence of variation across demographic subgroups in our LMIC samples, as shown in Appendix A Table 9. Women were generally less willing to accept the vaccine (average difference about 4.2 points, significant at *p* < .01). Respondents under age 24 and less educated respondents were marginally more willing to take the vaccine, but these differences were not statistically significant. The acceptance advantage among the less educated was most pronounced in Rwanda, Burkina Faso, and Uganda, and stands in contrast to the USA, where more educated respondents were significantly more willing to be vaccinated.

To better understand the reasoning behind vaccine acceptance, we asked those who were willing to take the vaccine why they would take it. We summarize these results in Table 3, with additional details in Appendix A Table 5. The reason most commonly given for vaccine acceptance across samples was personal protection against COVID-19 infection. The average across the LMIC samples was 91% (86–96%). In every individual study, this ranked as the most popular reason. In distant second place among LMIC respondents was family protection with an average of 36% (28– 43%). In comparison to protecting oneself and one’s family, protecting one’s community did not feature prominently among stated reasons for acceptance. These reasons do not vary substantially by age group, as shown in Table 11. Self-protection also ranked as the most common reason for taking the vaccine in Russia (76%, 74–78%) and the USA (94%, 92–95%).

Figure 2 summarizes the reasons given by respondents who said they were not willing to take a COVID-19 vaccine. Concern about side effects was the most frequently expressed reason for reluctance in our LMIC samples. This concern was particularly evident among African country samples. In studies Uganda 1 (85.1% 80.7–89.6%), Sierra Leone 2 (57.9%, 50.1–65.7%), Sierra Leone 1 (53.5% 47.1–59.9%) and Uganda 2 (47.3% 42.2–52.5%), more than half of respondents unwilling to take the vaccine cited worries about side effects. Respondents in Russia (36.8%, 35.2– 38.4%) and even more in the USA (79.3%, 74.6–84%), frequently reported this same concern.

**Figure 2:**
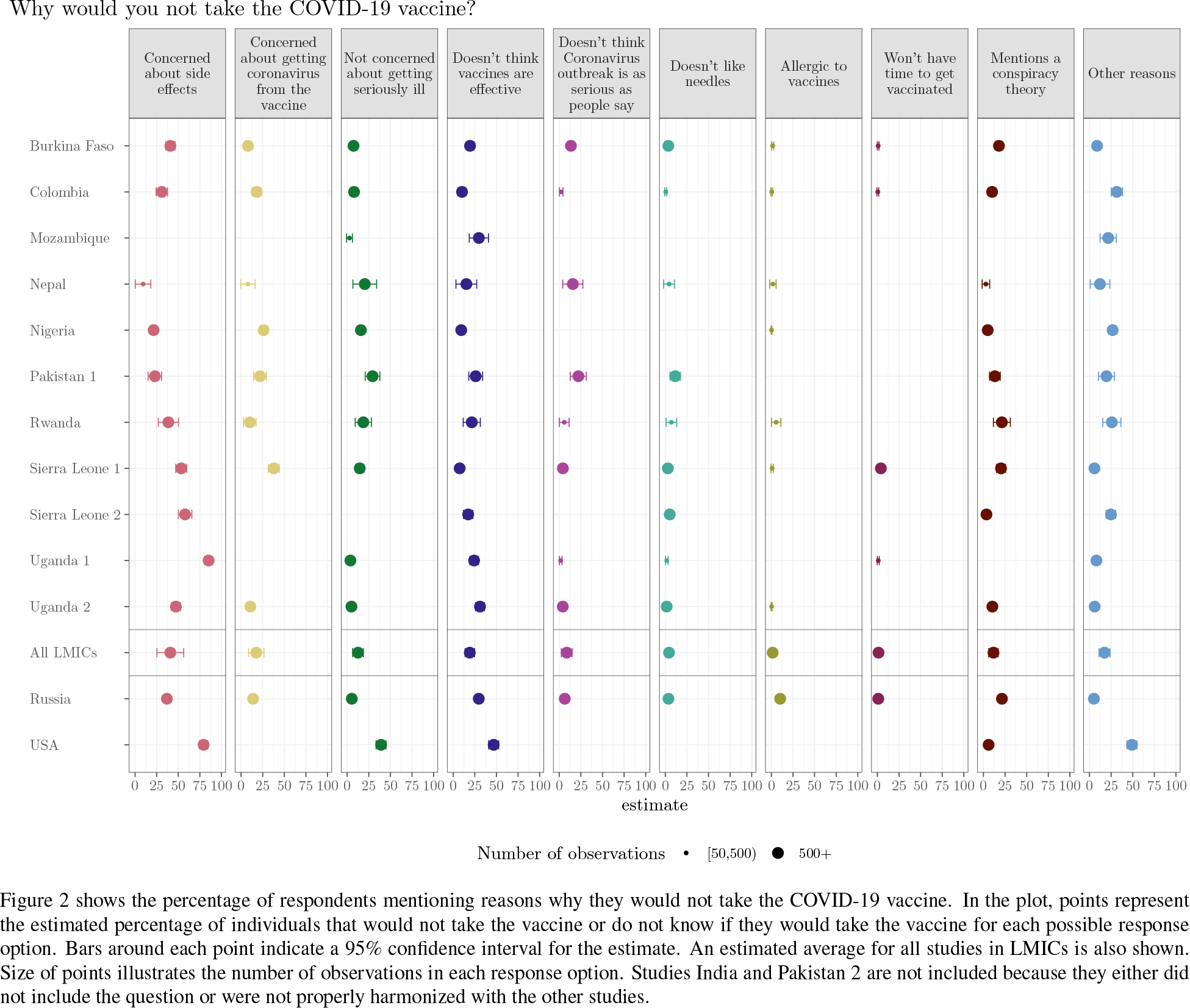
Reasons not to take the vaccine.

Study samples Uganda 2 (31%, 25.9–36.2%), Mozambique (29.7%, 18.6–40.8%) and Pakistan 1 (26%, 18–34%) showed relatively high levels of skepticism about vaccine effectiveness among hesitant respondents. This was also true in Russia (29.6%, 28.1–31.1%) and the USA (46.8%, 41–52.6%). In addition, hesitant respondents listed lack of concern about COVID-19 infection as a reason not to be vaccinated. Study samples USA (39.3% 33.5–45%), Pakistan 1 (29.4%, 20.9–37.9%) and Nepal (20.4% 6.7–34.1%) reported high rates of this answer among hesitant respondents.

In Figure 3 we report respondents’ most trusted source of guidance when deciding whether to take a COVID-19 vaccine. Results from Figure 3 are reproduced as Appendix A Table 8. Appendix B Table 15 presents a complete description of response recoding from individual studies.

**Figure 3:**
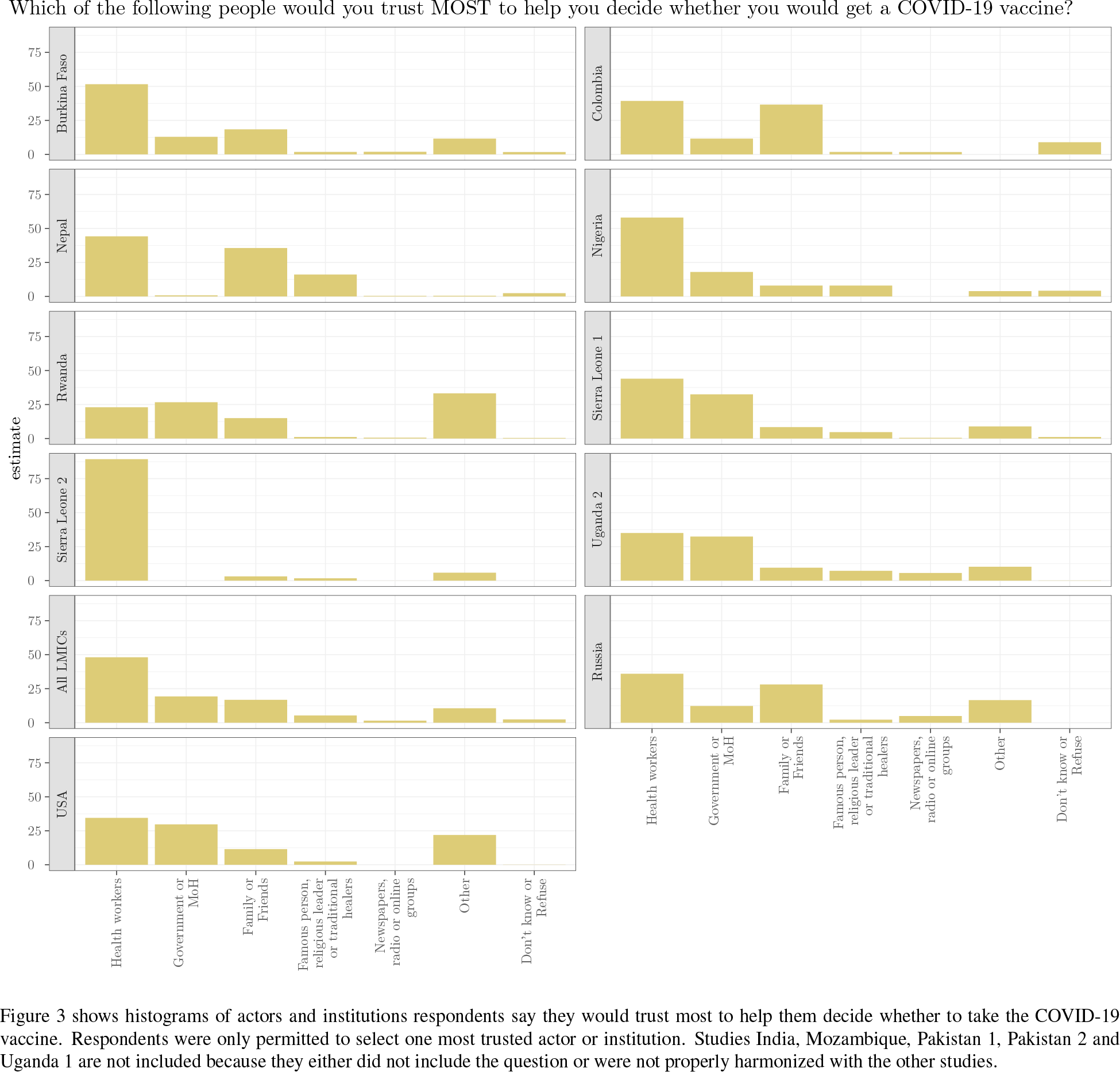
Trusted actors and institutions, broken down by expressed willingness to take a COVID-19 vaccine.

We find striking consistency in most trusted sources across studies. In all but one sample, including those from Russia and the USA, respondents identified the health system as the most trustworthy source to help them decide whether to take the COVID-19 vaccine. The average across LMICs was 48.1% (31.6–64.5%). Respondents in Sierra Leone 2 (89.3%, 87.2–91.5%), Nigeria (58%, 55.7– 60.2%) and Burkina Faso (51.6%, 48.5–54.8%) cited health workers most often. Sierra Leone had the highest trust in health workers and the Ministry of Health, potentially due to investments in public health following the 2014-2015 Ebola epidemic.^23^

The next most cited sources were family and/or friends in Colombia (36.6%, 33.5–39.7%), Nepal (35.6%, 32.9–38.3%), Russia (28.1%, 26.8–29.3%) and Burkina Faso (18.4%, 16–20.9%). Across the pooled samples, women were 3 percentage points more likely to rely on family and friends than male respondents, though this difference was not statistically significant (Figure 5 in Appendix D).

By contrast, in Sierra Leone 1 (32.5%, 29.7–35.4%), Uganda 2 (32.4%, 29.9–35%), USA (29.7%, 27–32.3%) and Nigeria (18%, 16.2–19.8%), government was the second most frequently cited. Religious leaders and celebrities were not seen as the top sources of guidance by many respondents in any sample other than Nepal, where many respondents most trusted famous people (16.1%, 13.3–18.9%).

Finally, we highlight two idiosyncratic, yet frequently mentioned, trusted sources of information in deciding whether to take a COVID-19 vaccine. In Rwanda, 34% of respondents would most trust “themselves” for guidance, the most frequent response in this sample. In the USA, 14% of respondents cited Joe Biden, then president-elect and therefore excluded from the “government” category, as their most trusted source.

## Discussion

To our knowledge, this is the first study focusing on COVID-19 vaccine attitudes in a large set of low-income and lower-middle-income countries. Our findings showed variable but broadly high levels of prospective COVID-19 vaccine acceptance across LMICs, using data from 44,260 respondents in 13 studies in ten LMICs in Africa, South Asia, and Latin America. Acceptance across these LMIC samples averaged 80.3%, ranging between 66.5% and 96.6%. The two benchmark countries, Russia and the USA, demonstrated lower COVID-19 vaccine acceptance, consistent with relatively lower pre-pandemic vaccine confidence.

Many metrics and indices measure vaccine acceptance and hesitancy globally.^24–26, 27^, Our surveys used measures employed in other COVID-19 vaccine acceptance studies^3, 6–11^ and recommended by the WHO Data for Action guidance,^28^ allowing for meaningful cross-study and cross-country comparisons. We measure trust in sources of information about COVID-19 vaccination using a measure similar to that used in the Vaccine Confidence Index (VCI), a widely used survey tool.^4^

Consistent with other studies, we find higher vaccine acceptance among men than women.^3, 7–10^ In contrast to studies focused primarily on higher-income countries, we find no consistently significant differences with respect to age^7, 9^ or education in our LMIC samples.

A key contribution of our study relative to the previous literature is its focus on the reasons why respondents express intentions to take (or refuse) a COVID-19 vaccine. Other work has highlighted appeals to altruistic behavior or other prosocial motivations to promote vaccine acceptance.^29^ Yet we find that the potential risks and benefits to personal well-being feature much more prominently in our respondents’ reasoning, suggesting that appeals about personal protection may be more effective in LMICs.

The most commonly stated reason for vaccine refusal was concern about safety (side effects). Around 86% of our surveys were conducted as reports from Phase 2 and 3 clinical trial data were emerging for the earliest commercially available vaccines, but prior to the first Emergency Use Authorization of any vaccine (Pfizer-BioNTech approved by the USA on December 11, 2020). Early trial data showed that severe adverse effects were extremely rare,^30–35^ occurring in fewer than 10% of people in clinical trials.^36^ Our respondents’ outsized concern about side effects could reflect the rapid pace of vaccine development,^37^ and limited information available about potential COVID-19 vaccine safety at the time of data collection. These concerns could also reflect worries about mild, yet common and transient side effects, such as fatigue, muscle pain, joint pain and headache.

Prominent media coverage of adverse events may exacerbate concerns about side effects.^38^ In particular, new information about rare but severe cases of thrombosis associated with the Astra-Zeneca vaccine that appeared after our data collection period could affect hesitancy levels. This is of particular relevance to LMICs, which are likely to rely on the Astra-Zeneca vaccine in their immunization campaigns, through initiatives such as COVAX.

Concerns about vaccine efficacy, averaging approximately 19.2% in the LMIC samples, may also reflect a lack of information about vaccines at the time of our surveys. Strikingly, even in low-information environments, respondents in our samples rarely cited conspiracy theories about ulterior motives on the part of corporations, politicians or the pharmaceutical industry, despite widespread popular fears about these issues in higher-income countries.^39^

An important limitation of our study is that our samples are not fully nationally representative. Phone surveys, while necessary during a global pandemic, do not include individuals who reside outside coverage areas, lack access to a cell phone, or do not respond to calls. In addition, as shown in Table 2, several studies focus on sub-populations of interest in pre-existing studies to which questions about COVID-19 vaccination were added. Particular care should be taken in any attempt to extrapolate to the national population level from these samples. Furthermore, we emphasize that our data are not representative of all LMICs. They represent a convenience sample of studies in countries in which our organizations could quickly mobilize coordinated data collection.

**Table 2:** Summary of studies sampling.

| Study | Date | Geographic scope | Sampling methodology | Survey modality | Weights |
| --- | --- | --- | --- | --- | --- |
| Burkina Faso | October to December 2020 | National | Random digit dialing (RDD) | Phone | Yes |
| Colombia | August 2020 | National | Random digit dialing (RDD) | Phone | Yes |
| India | June 2020 to January 2021 | Subnational, Slums in 2 cities | Representative sample of slum dwellers living in vicinity of a community toilet and located in Uttar Pradesh | Phone | Yes |
| Mozambique | October to November 2020 | Subnational, 2 cities | 1) Random sample in urban and periurban markets stratified by gender and type of establishment in Maputo; 2) Random sample representative of communities in the Cabo Delgado, stratified on urban, semiurban, and rural areas | Phone | No |
| Nepal | December 2020 | Subnational, 2 districts | Random sample of poor households from randomly selected villages in Kanchanpur | Phone | Yes |
| Nigeria | November to December 2020 | Subnational, 1 state | 1) Random sample of individuals in Kaduna; 2) Sample of phone numbers from a phone list of Kaduna state residents | Phone | No |
| Pakistan 1 | July to September 2020 | Subnational, 2 districts | Random sample of individuals in administrative police units in two districts of Punjab | Phone | Yes |
| Pakistan 2 | September to October 2020 | Subnational, 1 province | Random digit dialing (RDD) on a random sample of all numerically possible mobile phone numbers in the region of Punjab | Phone | No |
| Russia | November to December 2020 | Subnational, 61 regions | Sample recruited from the Russian online survey company OMI (Online Market Intelligence). Sampling targeted at having a minimum of respondents per region, as well as representation of age, gender and education groups. | Online | Yes |
| Rwanda | October to November 2020 | National | Random digit dialing (RDD) | Phone | Yes |
| Sierra Leone 1 | October 2020 | National | Random digit dialing (RDD) | Phone | Yes |
| Sierra Leone 2 | October 2020 to January 2021 | National | A random sample of households in 195 rural towns across all 14 districts of Sierra Leone | Phone | No |
| Uganda 1 | September to December 2020 | Subnational, 13 districts | Sample of women in households from semi-rural and rural villages across 13 districts in Uganda, selected according to the likelihood of having children | Phone | No |
| Uganda 2 | November to December 2020 | Subnational, 1 district | Random sample of households in Kampala | Phone | No |
| USA | December 2020 | National | Nation-wide sample of adult internet users recruited through the market research firm Lucid | Online | Yes |

**Table 3:** Reasons to take the vaccine.

| Study | N | Protection |  |  |
| --- | --- | --- | --- | --- |
|  |  | Self | Family | Community |
| Burkina Faso | 651 | 76<br>(73, 79) | 42<br>(38, 46) | 7<br>(5, 9) |
| Colombia | 756 | 91<br>(88, 93) | 23<br>(20, 26) | 12<br>(10, 14) |
| Mozambique | 768 | 83<br>(80, 86) | 32<br>(27, 38) | 4<br>(2, 5) |
| Nepal | 1341 | 96<br>(95, 98) | 34<br>(32, 37) | 20<br>(17, 22) |
| Nigeria | 1424 | 89<br>(88, 91) | 35<br>(33, 38) | 21<br>(19, 23) |
| Rwanda | 1152 | 98<br>(97, 99) | 26<br>(23, 28) | 11<br>(9, 13) |
| Sierra Leone 1 | 836 | 94<br>(92, 96) | 37<br>(34, 40) | 21<br>(18, 23) |
| Sierra Leone 2 | 1855 | 91<br>(88, 93) | 62<br>(57, 66) | 21<br>(16, 27) |
| Uganda 1 | 2885 | 96<br>(95, 97) | 36<br>(34, 38) | 9<br>(8, 10) |
| Uganda 2 | 1045 | 96<br>(95, 97) | 28<br>(25, 31) | 11<br>(9, 12) |
| All LMICs | . | 91<br>(86, 96) | 36<br>(28, 43) | 14<br>(9, 18) |
| Russia | 5887 | 76<br>(74, 78) | 69<br>(67, 71) | 41<br>(38, 43) |
| USA | 1313 | 94<br>(92, 95) | 92<br>(90, 94) | 89<br>(87, 91) |

Table 3 shows percentage of respondents mentioning reasons why they would take the Covid-19 vaccine. The number of observations and percentage corresponds only to people who would take the vaccine. Respondents in all countries could give more than one reason. A 95% confidence interval is shown between parentheses. Studies India, Pakistan 1 and Pakistan 2 are not included because they either did not include the question or were not properly harmonized with the other studies.

The expressed intentions to take a COVID-19 vaccine that we document in our LMIC samples, if translated into behavior, would meet or exceed the current herd immunity threshold for COVID-19 (estimated at 60-80%, based on the predominant variant in circulation in these countries).^40–42^ However, reported intent may not always convert vaccine uptake.^43^ The high salience of COVID-19 due to extensive media coverage may have increased reported intentions. Conversely, reports about side effects and risks associated with expedited vaccine development may have increased hesitancy. The fast-moving pandemic and vaccine development context may change perceptions about vaccines by the time they are widely available in LMICs.

Indeed, previous research on vaccine hesitancy has emphasized how concerns that arise surrounding vaccination campaigns are often case- and context-specific,^44^ making it difficult to predict exactly how COVID-19 vaccines will be received in any given setting. The lower COVID-19 vaccine acceptance rates we observe in Russia and the USA, for example, may reflect the politicization of this specific pandemic and vaccine development,^45–48^ in addition to generally greater vaccine skepticism.

Nonetheless, our findings suggest several concrete implications for policy relating to vaccine roll-out in LMICs. First and foremost, we document high levels of COVID-19 vaccine acceptance in LMICs compared to Russia and the USA. While global vaccine distribution has skewed heavily toward higher-income countries to date,^3^ our findings suggest that prioritizing distribution to LMICs is justified not only on equity grounds, but on the expectation of “better value” in maximizing global coverage more quickly.

Our findings also imply that, once vaccine distribution to LMICs begins in earnest, interventions should focus on converting positive intentions into action. Straightforward, low-cost nudges may be particularly effective in this regard. Two recent large-scale studies in the USA found that vaccination appointment reminder messages from healthcare providers increased influenza vaccine uptake.^49, 50^ Similar interventions have proven effective in increasing immunization in LMIC contexts. In Ghana and Kenya, vaccination reminders plus small cash incentives increased childhood immunization coverage.^51, 52^ Cash and in-kind incentives programs were also effective in Nigeria and India.^53, 54^

This recommendation is consistent with accepted frameworks, such as the WHO’s Behavioral and Social Drivers of vaccination (BeSD) model, which suggests leveraging favorable intentions through reminders and primes, and reducing access barriers when the vast majority of people intend to be vaccinated.^28, 55^ Particularly since COVID-19 vaccination may be more collectively than individually optimal, ease of access is critical to achieve high coverage.^56^

Our findings also suggest directions for the design and delivery of messaging to boost COVID-19 vaccine acceptance and uptake in LMICs. We highlight four potential implications. First, our data strongly support the view that respondents from LMICs prefer to follow the guidance of actors with the most relevant knowledge and expertise. We find high levels of trust in health workers, which suggests that social and behavioral change communication (SBCC) strategies engaging local health workers may be particularly effective in encouraging efficient and comprehensive vaccine uptake and combating remaining hesitancy.^46, 57^ Health workers have been the first group to receive the COVID-19 vaccine and are therefore best-positioned to share locally credible experiences of vaccination.^58^ While celebrities were not identified as a particularly trustworthy source for COVID-19 advice in our study, celebrity endorsements have proven effective in other contexts and may complement a strategy that primarily focuses on health workers.^59^

Second, our findings offer guidance on the specific content of vaccine messaging that is likely to be most persuasive. Hesitant respondents were most concerned about side effects and vaccine efficacy. This suggests that messaging should highlight the high efficacy rates of the COVID-19 vaccines currently on the market in reducing or eliminating disease, hospitalizations, and death, and communicate accurate information about potential side effects. Our data also suggest that messaging should emphasize the direct protective benefits of the vaccine to the adopter, since personal protection, rather than broader concerns about protecting public health, was the top reason expressed for vaccine-acceptance by our respondents.

Third, consistent with previous studies on COVID-19 vaccination^3, 7–10^ our study finds lower vaccine acceptance among women than men, suggesting that messaging strategies should focus on women. Recent work in Latin America on COVID-19 vaccine messaging found that the provision of basic information about the vaccines was particularly effective in persuading hesitant women.^60^

Finally, high coverage rates of existing vaccines, coupled with respondents’ reliance on friends and family as information sources, suggest that the general pro-vaccination stance of many LMIC citizens could be leveraged to increase uptake of COVID-19 vaccines as they become available. Social learning strategies and norm-setting are powerful drivers of behavior in many related sectors.^61^ Social signaling of positive attitudes toward and uptake of COVID-19 vaccines may also help shift social norms toward even greater immunization acceptance and two-dose completion in the community at large.^62^

## Methods

### Survey questions and sample construction

Survey data were collected between June 2020 and January 2021. Our main outcome measure is vaccine acceptance. Across studies, we asked respondents, “If a COVID-19 vaccine becomes available in [your country], would you take it?”. This measure aligns with widely reported COVID-19 vaccine acceptance measures.^3, 6–11^ If the respondent answered yes to this question, we followed up with the question, “Why would you take it? [the COVID-19 vaccine]”. If the respondent said they would not be willing to take the vaccine, we followed up with the question, “Why would you not take it? [the COVID-19 vaccine]”. Finally, regardless of their expressed willingness to take the vaccine, we asked about actors and institutions that would be most influential in their decision: “Which of the following people would you trust MOST to help you decide whether you would get a COVID-19 vaccine, if one becomes available?” following.^4^ To examine heterogeneity across demographic strata, we collected information about gender, age, and education. Slight variations in question wording and answer options across studies are documented in Appendix B.

Studies vary in terms of geographic scope, sampling methodology, and survey modality. Seven were national or nearly-national in scope. Studies from Burkina Faso, Colombia, Rwanda, and Sierra Leone (“Sierra Leone 1”) used nationally-representative samples of active mobile phone numbers reached through Random Digit Dialing (RDD). Studies in the USA and Russia were conducted online using quota samples obtained from private survey companies.

The remaining eight studies targeted sub-national populations. One study from Pakistan (“Pakistan 2”) used RDD in Punjab province. Respondents in Mozambique, Nigeria, Pakistan (“Pakistan 1”), Uganda (“Uganda 1”,“Uganda 2”), India, Nepal and Sierra Leone (“Sierra Leone 2”) were drawn from pre-existing studies to which COVID-19 vaccine questions were subsequently added. For example, Sierra Leone 2 has national coverage from a study on access to electricity and Uganda 1 sampled female caregivers of households in rural and semi-rural villages as part of a large ongoing cluster-RCT implemented across 13 districts.

Table 2 in Appendix A summarizes the geographic scope, sampling methodologies and survey modalities of all 15 studies. A detailed description of each study is included in Appendix C.

All surveys were conducted remotely to minimize in-person contact and comply with social distancing guidelines. Interviews were conducted by local staff in each country in local language(s). Surveying by phone made rapid, large-scale data collection possible. In two samples, the USA and Russia, surveys occurred via online polling. All surveys lasted approximately 15 to 40 minutes.

Taken together, we have data from 20,176 individuals from 10 LMICs and 24,084 from the USA and Russia, for a total of 44,260 respondents.

### Statistical Analysis

Vaccine acceptance was defined as the percentage of respondents who answered “yes” to the question, “If a COVID-19 vaccine becomes available in [country], would you take it?”. This was calculated combining all other answer options (“No”, “Don’t Know” and “Refuse”) into a single reference category. We estimated average acceptance for each individual sample via ordinary least squares (OLS) weighted by respective study population weights and robust standard errors clustered at the level relevant for the sample.

In addition to study-level estimates, we combined data from all studies other than the USA and Russia to calculate an aggregate “All LMIC studies” estimate. For these analyses, we estimated average acceptance by OLS with weights for each study normalized such that the total weight given to observations was constant across studies. Robust standard errors for these analyses were clustered at the study level.

We note the core results would be virtually unchanged at 80.8% (74.5–87.1) rather than 80.3% (74.9 -85.6) using countries rather than studies as groups in the pooled analysis, that is if we set weights so that the sum of weights in each country (rather than in each study) sum to a constant and cluster standard errors at the country level (rather than the study level).

In this combined analysis, we also estimated the underlying heterogeneity of vaccine acceptance across studies using the between studies variance estimator *τ*^2^ from a random effects model.

We conducted subgroup analyses by gender, age and education level and reported differences between groups. For age, we selected cut-offs below 25, between age 25 and 54, and above 55 years old, closely following the age breakdown proposed by recent work on COVID-19 vaccine acceptance.^11^ However, the lower life expectancy (63 years on average)^63^ and younger-skewing populations (only 5% of the population is above 65 years old)^64^ of low-income countries in particular, precluded further disaggregation at the upper end of the age distribution. For education, we divided the sample between respondents who had completed secondary school and those that had not. We defined these two groups to reflect broader schooling trends in LMICs, where out of every 100 students entering primary education, 61% complete lower secondary education.^65^ The subgroup analyses estimates are calculated in exactly the same way as the overall acceptance rate—with weights again normalized to sum to a constant within each study—with the exception that the subsample used in the analysis is limited to those respondents fitting each demographic group.

We then investigated stated reasons for COVID-19 vaccine acceptance and hesitancy, and the types of actors respondents would trust most when making the decision about whether to take a COVID-19 vaccine. We report estimates of agreement with reasons for vaccine acceptance/hesitancy and trust in actors for individual studies and for the “All LMICs” group, which includes all study samples except Russia and the USA. Estimates were calculated with the same procedure as above, varying only the quantity of interest; i.e. one model is run for each reason why a respondent would (or would not) take the vaccine and each trusted actor.

## Data Availability Statement

Individual participant data (de-identified) that underlie the results reported in this article, are available without restrictions at https://github.com/wzb-ipi/covid_vaccines_nmed.

## Code Availability

All code has been deposited into the publicly available GitHub repository at https://github.com/wzb-ipi/covid_vaccines_nmed. The code and output for all analyses can be easily inspected at https://wzb-ipi.github.io/covid_vaccines_nmed/replication.html.

## Data Availability

Code and output of the analysis can be consulted here
https://wzb-ipi.github.io/covid_vaccines/replication.html.

## Acknowledgements

This work would not have been possible without the hard work and ingenuity of many. For valuable contributions to data collection during the challenging context of the global pandemic, we would like to thank:

Burkina Faso: Béchir Wendemi Ouédraogo (M&E Associate), Achille Mignondo Tchibozo (Research Manager) and Touba Bakary Pare (Senior Field Manager) at IPA Burkina Faso in Ouagadougou for their work on the RECOVR (Research for Effective Covid Response) survey from which the questions used in this study were drawn.

Colombia: Margarita Rosa Cabra Gacia (Senior Research Associate), Laura Polanco, María Juliana Otálora (Research Analyst), Sofía Jaramillo (Research Manager) and Mery Galindo (Field Coordinator) at IPA Colombia in Bogotá for their work on the RECOVR (Research for Effective Covid Response) survey from which the questions used in this study were drawn.

India: Bhartendu Trivedi, Yashashvi Singh, Tatheer Fatima and the field team of Morsel Research and Development.

Mozambique: Alberto Arlindo, Imamo Mussa, Ismail Mussa, Frederica Mendonça and Simão Paiva from NOVAFRICA Mozambique office.

Nepal: Arjun Kharel and field staff at the Centre for the Study of Labour and Mobility (CESLAM) in Kathmandu.

Nigeria: Opeyemi Adeojo (Department of Sociology, University of Lagos), Deborah Anigo, Isaac Obara, Segun Fayomi, and Aishwarya Kumar at the Busara Center for Behavioral Economics, and Sarah Ryan (University of Illinois at Chicago) for research assistance.

Pakistan 1: Ahsan Tariq and Shan e Muhammad Malik at the Institute for Development and Economic Alternatives (IDEAS) in Lahore as survey wing leads and Hamid Ali Tiwana for research assistance.

Pakistan 2: Staff and the internal survey firm at the Centre for Economic Research Pakistan (CERP).

Rwanda: Jean Leodomir Habarimana Mfura (Research and Policy Coordinator), Jean Aime Nsabimana and Gisele Manirabaruta (Field Manager) at IPA Rwanda in Kigali for their work on the RECOVR (Research for Effective Covid Response) survey from which the questions used in this study were drawn.

Russia: Staff at Online Market Intelligence Survey Agency, Kirill Chmel (HSE University) and Vladimir Zabolotsky (HSE University and University of Bologna) for their intellectual contributions and research assistance.

Sierra Leone 1: Filippo Cuccaro (Research Coordinator) and Fatoma Momoh (Senior Field Manager) at IPA Sierra Leone in Freetown for their work on the RECOVR (Research for Effective Covid Response) survey from which the questions used in this study were drawn.

Sierra Leone 2: Sellu Kallon (Field Manager), Junisa Nabieu (Field Supervisor), Momoh Flee (Field Supervisor), Hamid Kai, Mohamed Lamin (Field Supervisor) and all enumerators at IGC-Sierra Leone Freetown office and Wageningen University’s Project “Tracking the Economic Consequences of COVID-19 in Sierra Leone”.

Uganda 1: Staff at IPA Uganda, especially Sarene Shaked and Gloria Ayesiga, for their intellectual contributions, research assistance, and support throughout the Uganda 1 study.

Uganda 2: Co-PIs Ana Garcia Hernandez, Leah Rosenzweig, Lily Tsai, and Elly Atuhumuza, Martin Atyera, and their team of survey enumerators and support staff at IPA Uganda.

United States: Staff at Lucid (survey firm) for distributing the survey and providing the online survey sample.

IGC thanks their resident country teams in Sierra Leone, Uganda, Pakistan, and Mozambique for their continued guidance, coordination of activities with researchers and direct engagement and dissemination activities with national policymakers:

- Herbert McLeod (Country Director) and Abou Bakarr Kamara (Country Economist) at IGC Sierra Leone
- Richard Newfarmer (Country Director), Priya Manwaring (Country Economist), and Jakob Rauschendorfer (Country Economist) at IGC - Uganda
- Ijaz Nabi (Country Director), Hina Shaikh (Country Economist), and Usman Naeem (Country Economist) at IGC - Pakistan
- Claudio Frischtak (Country Director) and Egas Daniel (Country Economist) at IGC - Mozambique

IPA thanks staff from its global offices for their research assistance and support throughout the RECOVR survey. Hugo Salas (IPA Mexico) contributed to data analysis for Burkina Faso, Rwanda and Sierra Leone. Margarita Rosa Cabra Garcia also contributed to data analysis for Colombia. At IPA New York and Washington, DC, Michael Rosenbaum and Savanna Henderson provided research assistance.

NOVAFRICA thanks Cátia Batista (NOVA SBE), Sandra Sequeira (LSE) and Inês Vilela (Royal Holloway, University of London and NOVAFRICA) for their intellectual contributions.

Centre for Economic Research Pakistan (CERP) thanks Javaeria Qureshi (University of Illinois Chicago), Taimur Shah (CERP), and Basit Zafar (University of Michigan) for their contributions to the Economic Vulnerability Assessment (EVA) project from which the questions used in the Pakistan 2 study were drawn.

## Contributors

JSo, SW, NMe, AS, NMc, GS, MV and AM are co-first authors. DK, MC, MT, MH, AMM and SO are co-last authors. AMM and SO are also the corresponding authors. DK, AMM, MT, NMe, MC, and MV conceived of the study and provided overall guidance. SAb and NMe led the literature search, with input from AS, NMc, SW, AMM, AM and JSo. SW, NMe, AS, NMc, MV, GS, AA, SA, BA, AB, EB, CMB, AC, EC, MF, AG, AK, SK, RL, MBN, MP, JSh, JSv, PV, LB, BZ, MC, SAs, AC, AF, AH, MC, MT, and MH oversaw data collection as part of other research efforts. SW, NMe, and MT coordinated the project across study samples. The following verified the underlying data for individual study samples: EC (Burkina Faso, Colombia, Rwanda, Sierra Leone 1), BA and AB (India), AS and RL (Nigeria), AG, JSv and MBN (Uganda 1), CMB and MH (Uganda 2), NMe and MV (Sierra Leone 2), GS (Russia), MF (Mozambique), AF and JSh (Pakistan 1), SAs (Pakistan 2), CV (Nepal), and NMc (USA). JSo, GS, MH and SA collated and processed all datasets used for the analysis. NMe, MH, AMM, JSo, GS, SW, AS, EC, EB, MT, MV and NMc did the data interpretation with guidance from SO and AM. JSo, GS, EC and MH verified final datasets and analysis. JSo and GS did the data analysis and produced output figures with input from MH, AMM, DK, SW, EC, MV, NMe and MT. MH supervised the data analysis. JSo, SW, NMe, AMM, AS, NMc and MV wrote the first draft of the manuscript, with guidance from AM and SO. JSo, SW, NMe, AS, NMc, MV, SAb, EB, MP, JSh, PV, BZ, MC, MT, MH, AMM and SO revised the manuscript. All authors approved the final version of the manuscript. All authors had full access to all the data used in this study and had final responsibility for the decision to submit for publication.

## Appendix A: Supplementary tables, figures and results

**Table 4:** If a COVID-19 vaccine becomes available in [country], would you take it? Disaggregated by subgroups.

| Country | Average acceptability | Gender |  | Education |  | Age |  |  |
| --- | --- | --- | --- | --- | --- | --- | --- | --- |
|  |  | Female | Male | > Secondary | Up to Secondary | <25 | 25-54 | 55+ |
| Burkina Faso | 66.5<br>(63.5, 69.5) | 62.1<br>(56.3, 67.9) | 68.4<br>(65.0, 71.9) | 60.8<br>(55.9, 65.8) | 70.1<br>(66.4, 73.8) | 76.0<br>(58.0, 94.0) | 63.2<br>(53.0, 73.4) | .<br>. |
| Colombia | 74.9<br>(72.2, 77.6) | 73.5<br>(70.1, 77.0) | 77.3<br>(73.0, 81.7) | 78.1<br>(73.6, 82.5) | 73.4<br>(70.1, 76.8) | 75.4<br>(67.5, 83.4) | 74.2<br>(70.2, 78.3) | 73.8<br>(65.0, 82.6) |
| India | 84.3<br>(82.3, 86.3) | 82.4<br>(79.0, 85.8) | 84.7<br>(82.5, 87.0) | 87.8<br>(81.1, 94.6) | 85.9<br>(82.5, 89.2) | 77.6<br>(71.6, 83.6) | 85.4<br>(83.4, 87.3) | 83.1<br>(77.6, 88.5) |
| Mozambique | 89.1<br>(86.5, 91.7) | 86.2<br>(82.5, 90.0) | 91.3<br>(88.4, 94.1) | 86.1<br>(81.8, 90.4) | 89.7<br>(86.5, 92.8) | .<br>. | 88.3<br>(85.3, 91.2) | 91.7<br>(88.1, 95.4) |
| Nepal | 96.6<br>(95.5, 97.6) | 96.4<br>(94.6, 98.2) | 96.4<br>(95.1, 97.7) | .<br>. | .<br>. | 97.8<br>(95.7, 99.8) | 96.6<br>(95.2, 97.9) | 93.8<br>(90.4, 97.2) |
| Nigeria | 76.2<br>(74.3, 78.2) | 74.9<br>(71.7, 78.1) | 77.0<br>(74.6, 79.4) | .<br>. | .<br>. | 69.0<br>(63.3, 74.7) | 77.6<br>(75.5, 79.7) | 74.7<br>(65.8, 83.6) |
| Pakistan 1 | 76.1<br>(70.0, 82.3) | 72.2<br>(65.6, 78.8) | 80.1<br>(73.8, 86.4) | 83.6<br>(76.6, 90.5) | 74.0<br>(67.4, 80.5) | 86.3<br>(78.4, 94.1) | 75.6<br>(69.3, 81.8) | 80.8<br>(64.9, 96.7) |
| Pakistan 2 | 66.5<br>(64.1, 68.9) | .<br>. | .<br>. | 71.4<br>(67.3, 75.5) | 64.2<br>(61.2, 67.1) | .<br>. | .<br>. | .<br>. |
| Rwanda | 84.9<br>(82.9, 86.8) | 79.4<br>(75.8, 83.0) | 88.0<br>(85.8, 90.2) | 71.4<br>(65.5, 77.2) | 87.7<br>(85.8, 89.7) | 88.1<br>(85.0, 91.1) | 83.8<br>(81.3, 86.3) | 73.3<br>(59.2, 87.3) |
| Sierra Leone 1 | 78.0<br>(75.5, 80.5) | 74.1<br>(69.5, 78.7) | 80.1<br>(77.2, 83.1) | 74.4<br>(70.1, 78.7) | 80.2<br>(77.0, 83.3) | 78.0<br>(72.4, 83.6) | 78.3<br>(75.4, 81.1) | 74.0<br>(61.4, 86.6) |
| Sierra Leone 2 | 87.9<br>(86.2, 89.6) | 88.6<br>(85.7, 91.5) | 87.7<br>(85.9, 89.5) | 88.8<br>(85.0, 92.5) | 87.8<br>(86.0, 89.6) | 82.9<br>(73.9, 91.9) | 87.6<br>(85.6, 89.5) | 90.0<br>(87.4, 92.6) |
| Uganda 1 | 85.8<br>(84.4, 87.2) | 85.8<br>(84.4, 87.2) | .<br>. | 80.1<br>(74.4, 85.9) | 84.8<br>(83.2, 86.5) | 85.5<br>(82.7, 88.4) | 85.9<br>(84.3, 87.4) | .<br>. |
| Uganda 2 | 76.5<br>(74.3, 78.7) | 74.9<br>(71.5, 78.3) | 78.0<br>(75.2, 80.9) | 68.6<br>(64.3, 72.9) | 79.8<br>(77.3, 82.2) | 76.5<br>(71.0, 82.1) | 77.0<br>(74.4, 79.6) | 73.7<br>(67.3, 80.0) |
| All LMICs | 80.3<br>(74.9, 85.6) | 79.2<br>(73.4, 85.0) | 82.6<br>(77.4, 87.9) | 77.4<br>(71.4, 83.4) | 79.8<br>(74.1, 85.4) | 82.8<br>(76.9, 88.7) | 81.1<br>(75.6, 86.6) | 79.1<br>(72.5, 85.7) |
| Russia | 30.4<br>(29.1, 31.7) | 22.6<br>(20.9, 24.2) | 38.5<br>(36.5, 40.5) | 31.0<br>(29.6, 32.5) | 29.6<br>(27.3, 32.0) | 33.5<br>(29.2, 37.7) | 27.6<br>(26.2, 28.9) | 40.0<br>(35.9, 44.0) |
| USA | 64.6<br>(61.8, 67.3) | 56.1<br>(52.1, 60.1) | 73.4<br>(69.8, 76.9) | 72.3<br>(69.5, 75.0) | 51.5<br>(46.0, 57.0) | 51.0<br>(43.5, 58.6) | 64.9<br>(61.1, 68.7) | 69.4<br>(64.8, 73.9) |

**Table 5:**
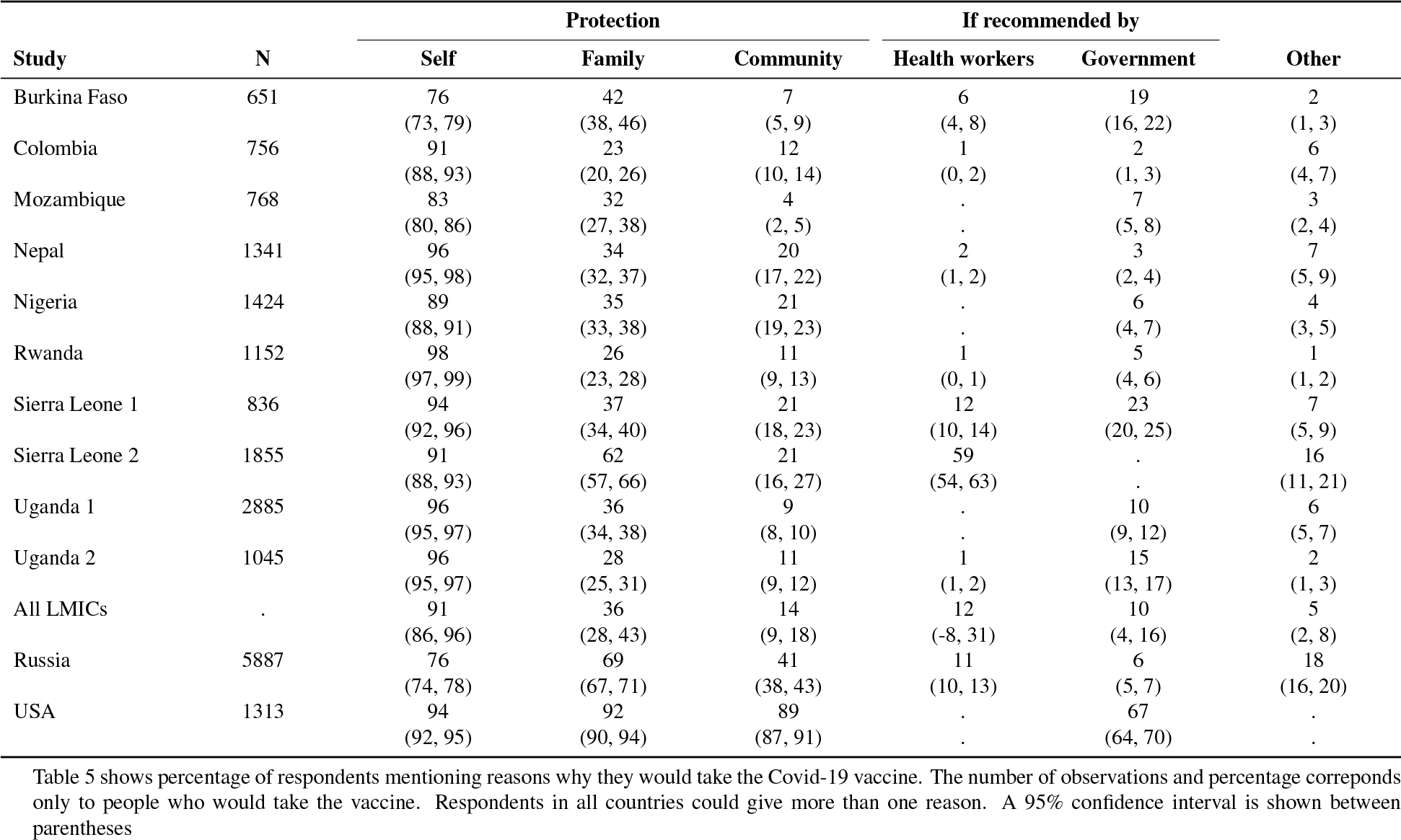
Reasons to take the vaccine- all categories.

**Table 6:** Reasons to take the vaccine- by age groups.

| Study | Self |  |  | Family |  |  | Community |  |  |
| --- | --- | --- | --- | --- | --- | --- | --- | --- | --- |
|  | <25 | 25-54 | 55+ | <25 | 25-54 | 55+ | <25 | 25-54 | 55+ |
| Burkina Faso | 77 | 59 | 100 | 26 | 64 | 66 | 11 | 2 | 0 |
| Conf. interval | (56, 99) | (46, 72) | (100, 100) | (4, 48) | (51, 77) | (-80, 211) | (-5, 26) | (-2, 5) | (0, 0) |
| n | 19 | 57 | 3 | 19 | 57 | 3 | 19 | 57 | 3 |
| Colombia | 91 | 91 | 90 | 26 | 26 | 16 | 12 | 13 | 14 |
| Conf. interval | (86, 97) | (88, 94) | (83, 97) | (17, 35) | (21, 31) | (8, 25) | (4, 20) | (9, 16) | (6, 22) |
| n | 90 | 349 | 73 | 90 | 349 | 73 | 90 | 349 | 73 |
| Mozambique | 62 | 84 | 80 | 50 | 32 | 34 | 12 | 4 | 2 |
| Conf. interval | (19, 106) | (81, 87) | (75, 86) | (5, 95) | (26, 38) | (27, 41) | (-17, 42) | (2, 6) | (0, 4) |
| n | 8 | 571 | 188 | 8 | 571 | 188 | 8 | 571 | 188 |
| Nepal | 97 | 97 | 92 | 31 | 36 | 27 | 15 | 20 | 19 |
| Conf. interval | (94, 100) | (96, 98) | (87, 97) | (25, 37) | (33, 39) | (19, 36) | (10, 20) | (17, 23) | (13, 25) |
| n | 225 | 890 | 162 | 225 | 890 | 162 | 225 | 890 | 162 |
| Nigeria | 91 | 89 | 94 | 31 | 36 | 31 | 22 | 21 | 21 |
| Conf. interval | (87, 95) | (87, 91) | (89, 100) | (25, 38) | (33, 39) | (20, 42) | (16, 29) | (18, 23) | (11, 31) |
| n | 178 | 1175 | 71 | 178 | 1175 | 71 | 178 | 1175 | 71 |
| Rwanda | 98 | 98 | 100 | 22 | 28 | 29 | 10 | 11 | 10 |
| Conf. interval | (97, 100) | (97, 99) | (100, 100) | (17, 26) | (24, 31) | (12, 46) | (7, 13) | (9, 14) | (-1, 21) |
| n | 389 | 732 | 31 | 389 | 732 | 31 | 389 | 732 | 31 |
| Sierra Leone 1 | 96 | 94 | 94 | 36 | 38 | 27 | 24 | 20 | 22 |
| Conf. interval | (93, 99) | (92, 95) | (86, 102) | (29, 44) | (34, 41) | (12, 42) | (17, 31) | (16, 23) | (8, 36) |
| n | 167 | 632 | 37 | 167 | 632 | 37 | 167 | 632 | 37 |
| Sierra Leone 2 | 87 | 90 | 93 | 52 | 62 | 62 | 29 | 22 | 18 |
| Conf. interval | (78, 97) | (88, 92) | (89, 97) | (39, 66) | (58, 67) | (56, 67) | (16, 42) | (16, 28) | (12, 25) |
| n | 63 | 1376 | 396 | 63 | 1376 | 396 | 63 | 1376 | 396 |
| Uganda 1 | 96 | 96 | . | 34 | 36 | . | 9 | 9 | . |
| Conf. interval | (95, 98) | (96, 97) | . | (30, 39) | (34, 39) | . | (6, 11) | (8, 11) | . |
| n | 526 | 2218 | . | 526 | 2218 | . | 526 | 2218 | . |
| Uganda 2 | 97 | 96 | 97 | 20 | 30 | 28 | 8 | 11 | 13 |
| Conf. interval | (94, 99) | (95, 97) | (94, 100) | (14, 26) | (27, 33) | (21, 36) | (4, 11) | (9, 13) | (7, 19) |
| n | 173 | 749 | 123 | 173 | 749 | 123 | 173 | 749 | 123 |
| All LMICs | 89 | 89 | 93 | 33 | 39 | 36 | 15 | 13 | 13 |
| Conf. interval | (81, 97) | (81, 98) | (89, 98) | (25, 41) | (29, 48) | (23, 48) | (10, 20) | (8, 18) | (7, 19) |
| n | 1838 | 8749 | 1084 | 1838 | 8749 | 1084 | 1838 | 8749 | 1084 |
| Russia | 67 | 76 | 81 | 74 | 68 | 68 | 46 | 40 | 38 |
| Conf. interval | (59, 74) | (73, 78) | (76, 87) | (68, 81) | (66, 71) | (62, 74) | (38, 54) | (38, 43) | (32, 44) |
| n | 552 | 5108 | 227 | 552 | 5108 | 227 | 552 | 5108 | 227 |
| USA | 92 | 91 | 97 | 89 | 91 | 94 | 90 | 89 | 89 |
| Conf. interval | (88, 96) | (89, 94) | (95, 99) | (83, 95) | (88, 93) | (91, 97) | (85, 95) | (86, 92) | (85, 93) |
| n | 153 | 687 | 473 | 153 | 687 | 473 | 153 | 687 | 473 |

Table 6 shows percentage of respondents mentioning reasons why they would take the Covid-19 vaccine by age groups. The number of observations and percentage corresponds only to people who would take the vaccine. Respondents in all countries could give more than one reason. A 95% confidence interval is shown between parentheses

**Table 7:**
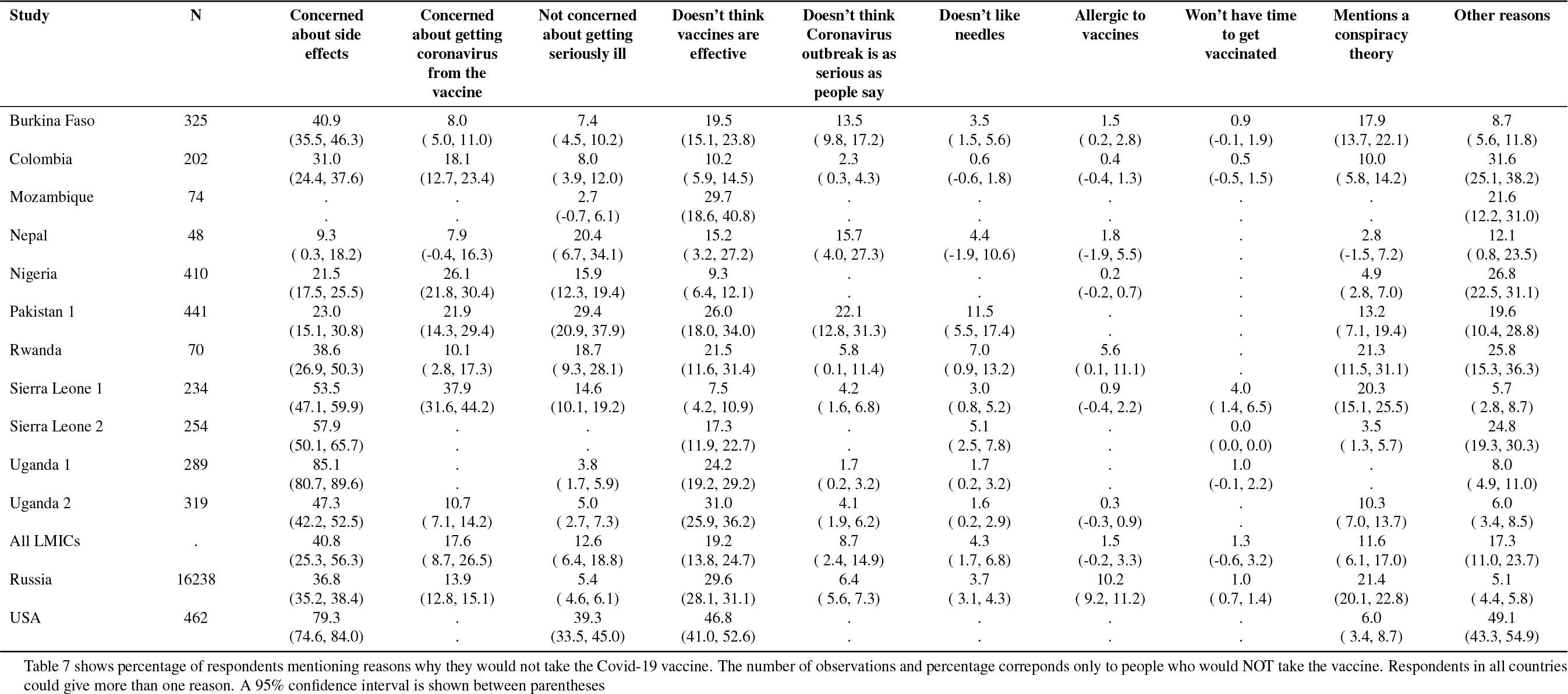
Reasons not to take the vaccine.

**Table 8:**
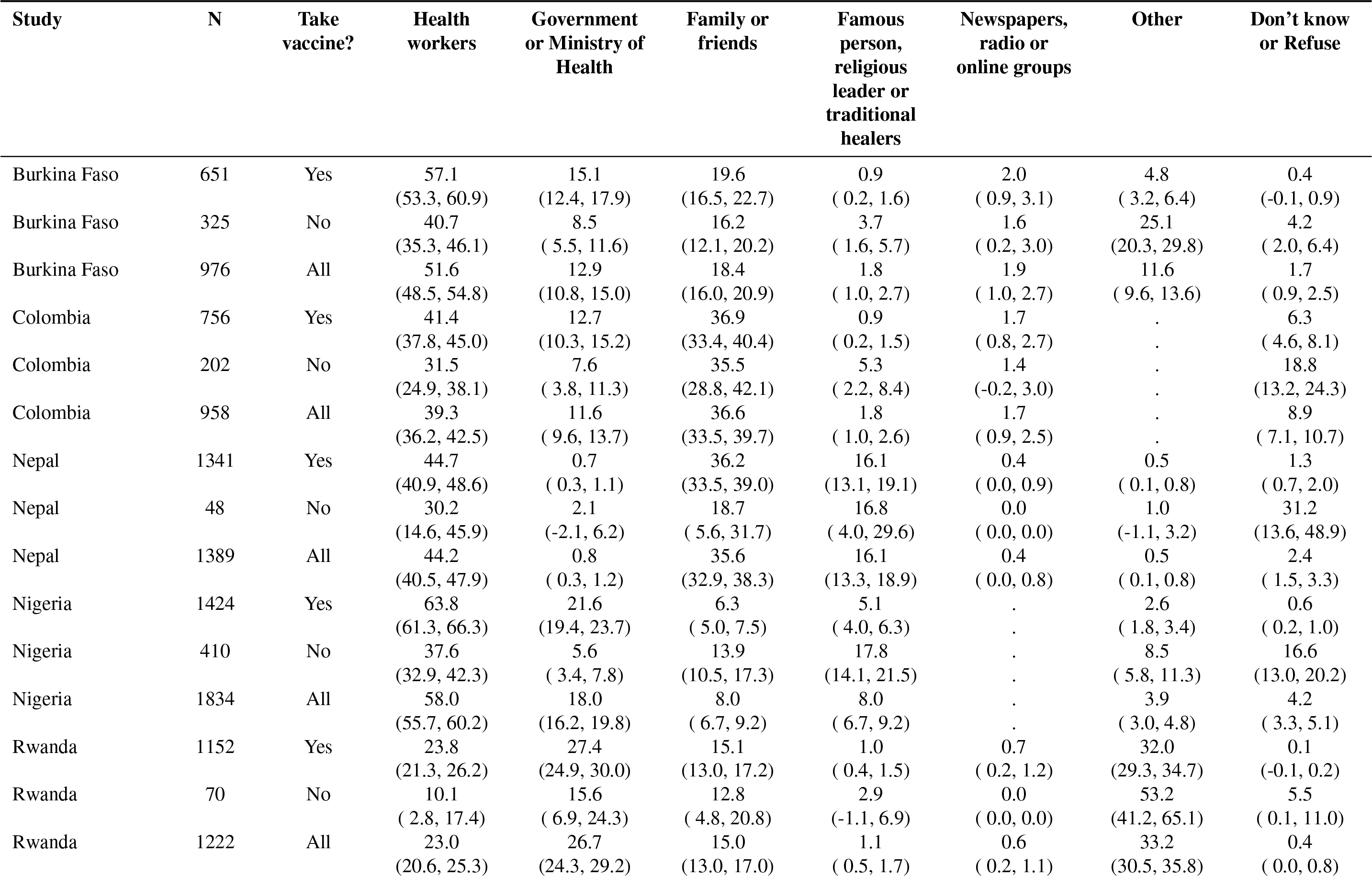
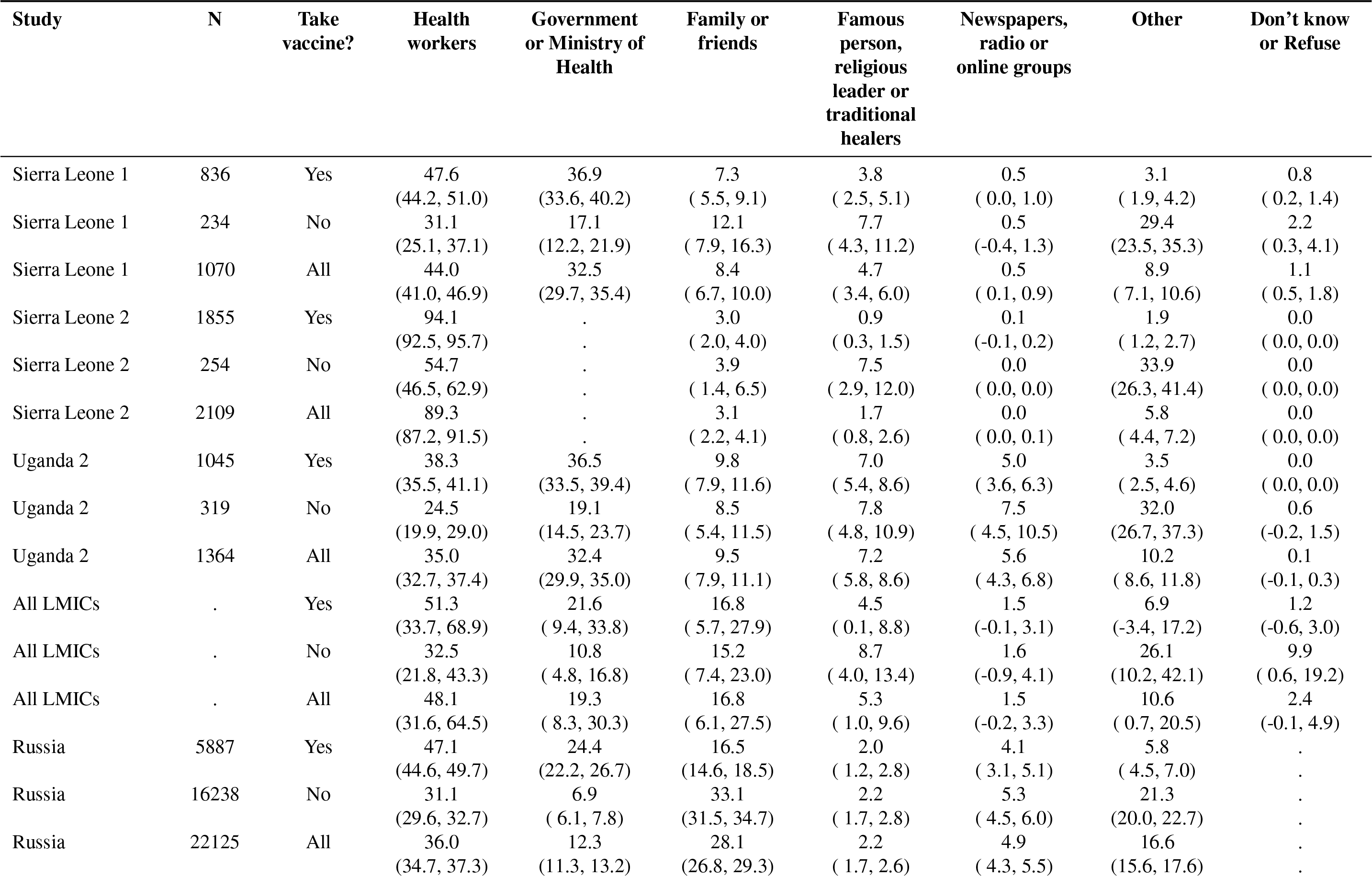
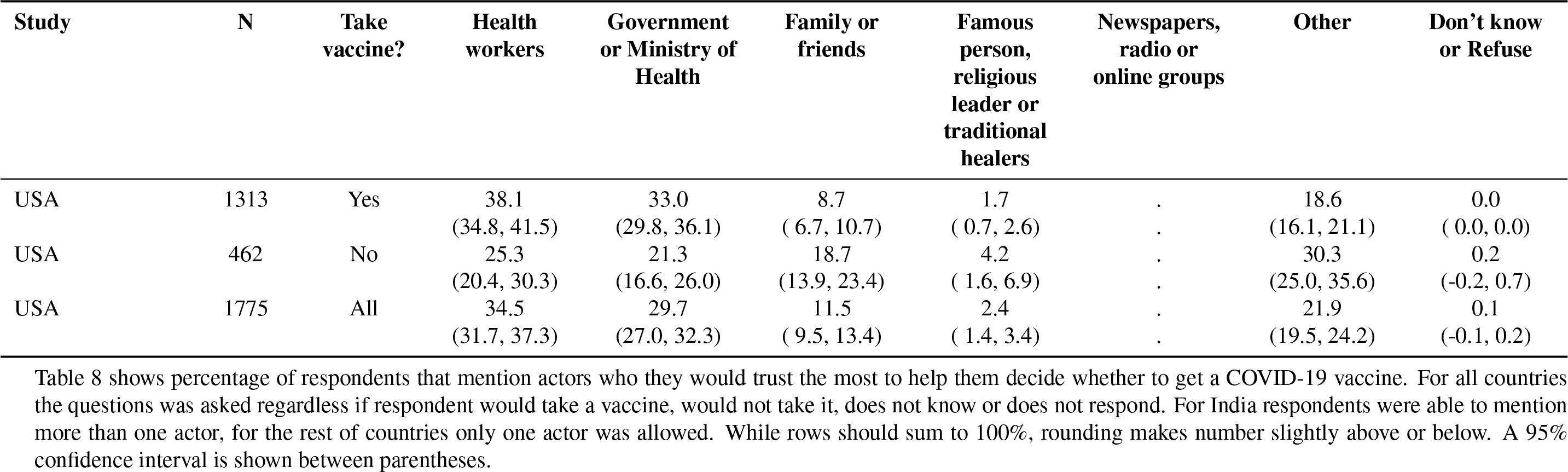
COVID-19 Vaccination Decision-making: most trusted source.

**Figure 4:**
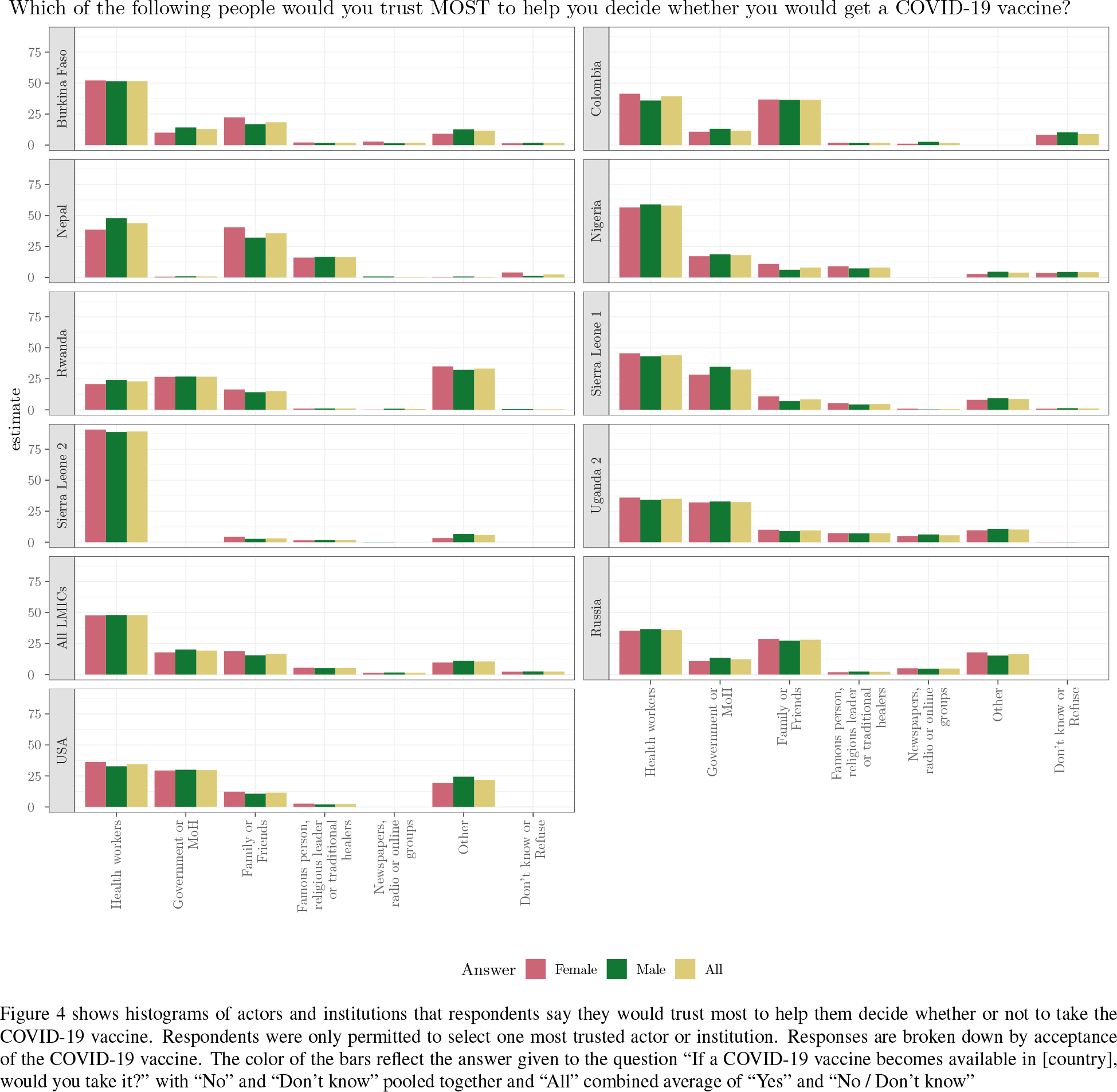
Trusted actors and institutions, broken down by gender.

**Figure 5:**
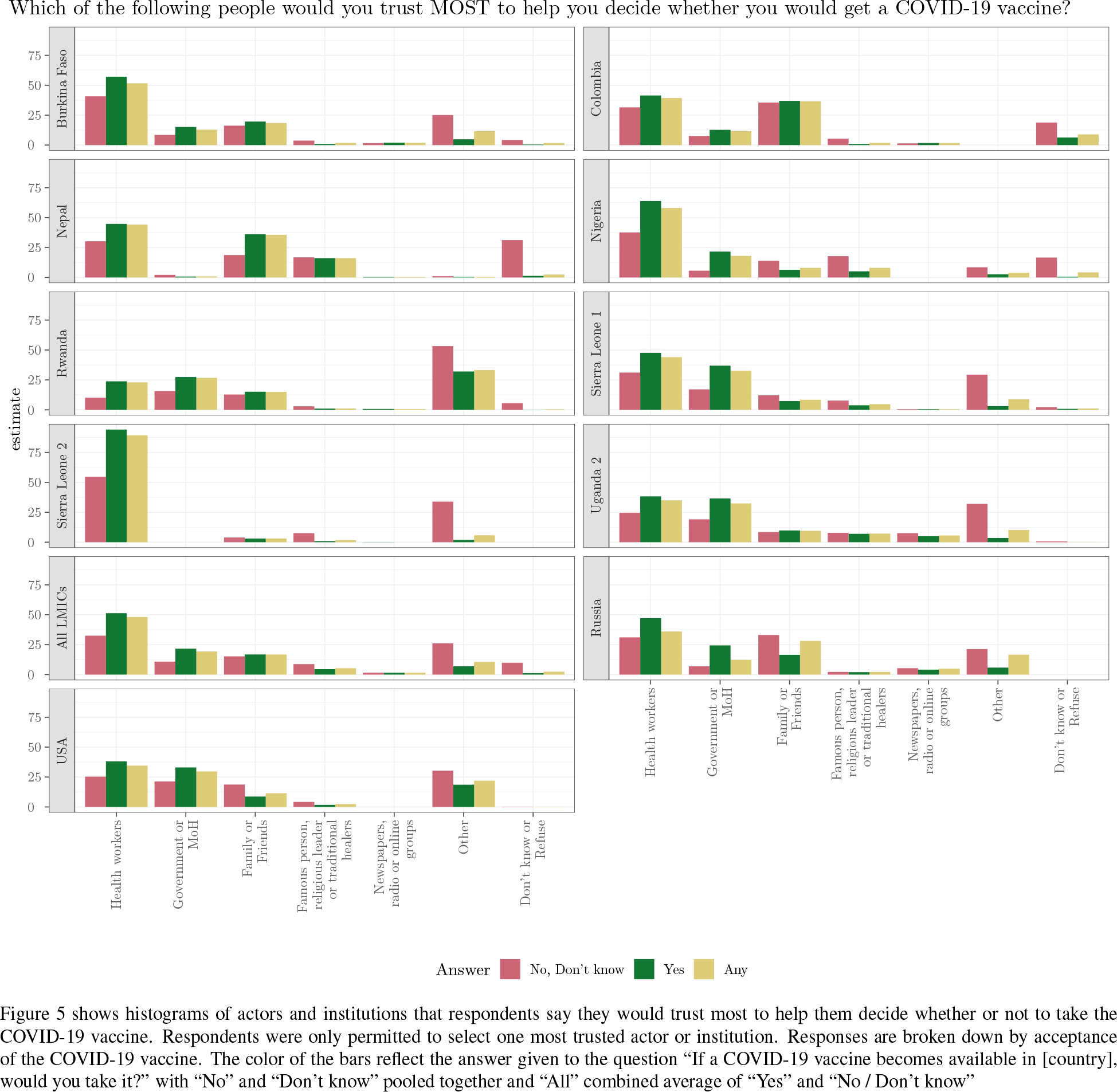
Trusted actors and institutions, broken down by vaccine acceptance.

**Table 9:** Differences in means.

| **Estimate** | **Std.error** | **P.value** | **Degrees of freedom** | **Baseline category** | **Variable** |
| --- | --- | --- | --- | --- | --- |
| 0.04 | 0.01 | 0.00 | 10 | Male | Gender (Female) |
| -0.02 | 0.02 | 0.43 | 10 | <25 | Age (25-54) |
| -0.02 | 0.02 | 0.36 | 10 | <25 | Age (55+) |
| 0.02 | 0.03 | 0.38 | 10 | Up to secondary | Education (Secondary +) |

Table 9 shows the results of subgroup mean differences. Subgroup differences were generated considering only LMICs. p-values come from a two-sided t-test from a linear regression.

**Table 10:** Observations and missingness patterns.

| Country | N obs | Take vaccine | Gender | Education | Age |
| --- | --- | --- | --- | --- | --- |
| Burkina Faso | 977 | 99.90 | 100.00 | 100.00 | 12.28 |
| Colombia | 1,012 | 94.66 | 100.00 | 99.90 | 68.18 |
| India | 1,680 | 100.00 | 100.00 | 20.24 | 100.00 |
| Mozambique | 862 | 97.68 | 100.00 | 96.06 | 99.77 |
| Nepal | 1,389 | 100.00 | 95.32 | 0.00 | 95.32 |
| Nigeria | 1,868 | 98.18 | 100.00 | 0.00 | 100.00 |
| Pakistan 1 | 1,633 | 98.96 | 99.76 | 99.27 | 100.00 |
| Pakistan 2 | 1,492 | 99.87 | 0.00 | 100.00 | 0.00 |
| Russia | 22,125 | 100.00 | 100.00 | 100.00 | 100.00 |
| Rwanda | 1,355 | 90.18 | 100.00 | 100.00 | 100.00 |
| Sierra Leone 1 | 1,070 | 100.00 | 100.00 | 97.01 | 100.00 |
| Sierra Leone 2 | 2,110 | 99.95 | 100.00 | 100.00 | 98.91 |
| Uganda 1 | 3,362 | 94.41 | 100.00 | 81.47 | 95.12 |
| Uganda 2 | 1,366 | 99.85 | 100.00 | 100.00 | 100.00 |
| USA | 1,959 | 90.61 | 100.00 | 100.00 | 100.00 |

Table 10 show the proportion of observations that are not missing values for each variable included in Figure 1.

**Table 11:**
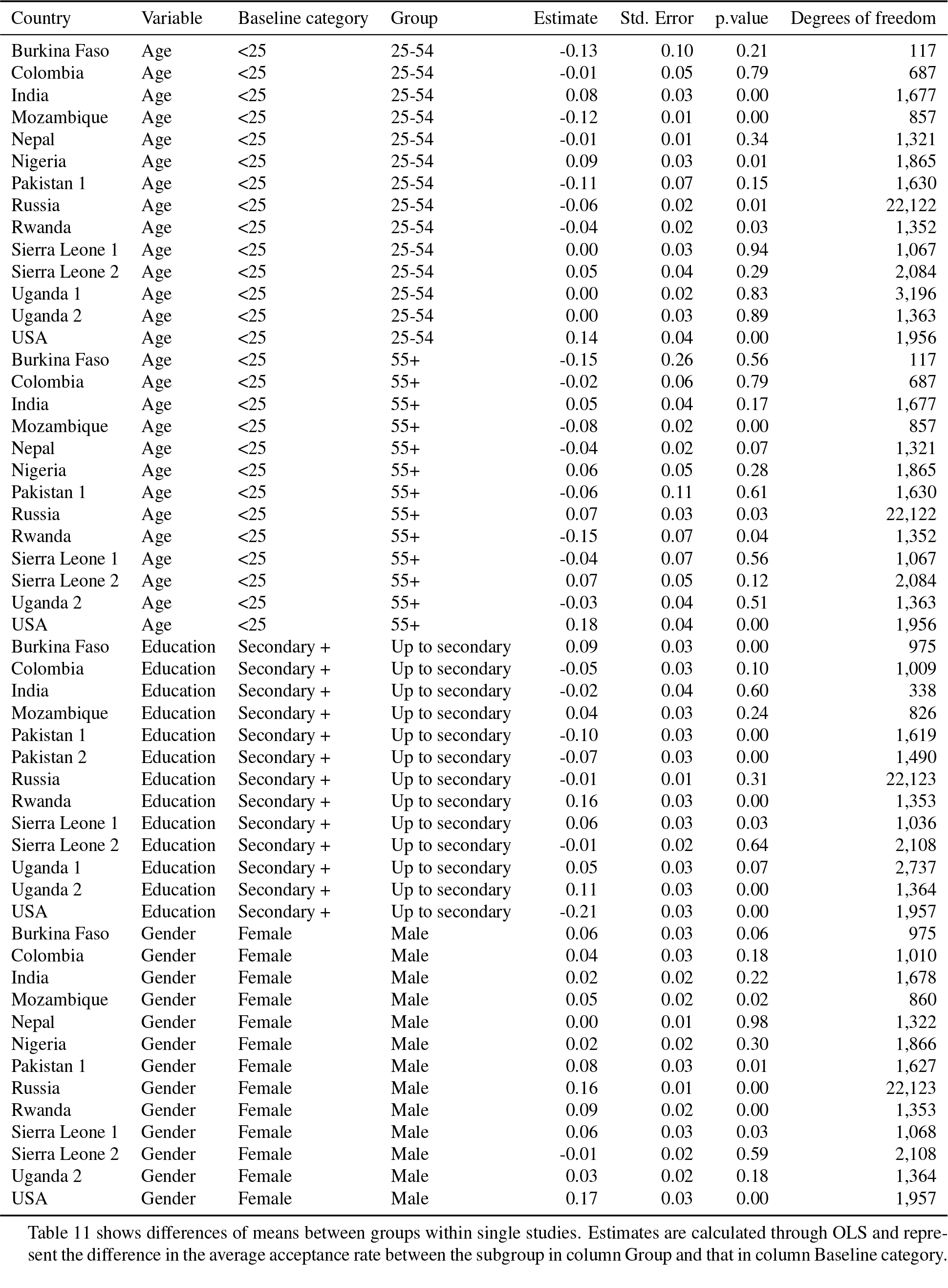
Differences between groups within studies.

| Country | Variable | Baseline category | Group | Estimate | Std. Error | p.value | Degrees of freedom |
| --- | --- | --- | --- | --- | --- | --- | --- |
| Burkina Faso | Age | <25 | 25-54 | -0.13 | 0.10 | 0.21 | 117 |
| Colombia | Age | <25 | 25-54 | -0.01 | 0.05 | 0.79 | 687 |
| India | Age | <25 | 25-54 | 0.08 | 0.03 | 0.00 | 1,677 |
| Mozambique | Age | <25 | 25-54 | -0.12 | 0.01 | 0.00 | 857 |
| Nepal | Age | <25 | 25-54 | -0.01 | 0.01 | 0.34 | 1,321 |
| Nigeria | Age | <25 | 25-54 | 0.09 | 0.03 | 0.01 | 1,865 |
| Pakistan 1 | Age | <25 | 25-54 | -0.11 | 0.07 | 0.15 | 1,630 |
| Russia | Age | <25 | 25-54 | -0.06 | 0.02 | 0.01 | 22,122 |
| Rwanda | Age | <25 | 25-54 | -0.04 | 0.02 | 0.03 | 1,352 |
| Sierra Leone 1 | Age | <25 | 25-54 | 0.00 | 0.03 | 0.94 | 1,067 |
| Sierra Leone 2 | Age | <25 | 25-54 | 0.05 | 0.04 | 0.29 | 2,084 |
| Uganda 1 | Age | <25 | 25-54 | 0.00 | 0.02 | 0.83 | 3,196 |
| Uganda 2 | Age | <25 | 25-54 | 0.00 | 0.03 | 0.89 | 1,363 |
| USA | Age | <25 | 25-54 | 0.14 | 0.04 | 0.00 | 1,956 |
| Burkina Faso | Age | <25 | 55+ | -0.15 | 0.26 | 0.56 | 117 |
| Colombia | Age | <25 | 55+ | -0.02 | 0.06 | 0.79 | 687 |
| India | Age | <25 | 55+ | 0.05 | 0.04 | 0.17 | 1,677 |
| Mozambique | Age | <25 | 55+ | -0.08 | 0.02 | 0.00 | 857 |
| Nepal | Age | <25 | 55+ | -0.04 | 0.02 | 0.07 | 1,321 |
| Nigeria | Age | <25 | 55+ | 0.06 | 0.05 | 0.28 | 1,865 |
| Pakistan 1 | Age | <25 | 55+ | -0.06 | 0.11 | 0.61 | 1,630 |
| Russia | Age | <25 | 55+ | 0.07 | 0.03 | 0.03 | 22,122 |
| Rwanda | Age | <25 | 55+ | -0.15 | 0.07 | 0.04 | 1,352 |
| Sierra Leone 1 | Age | <25 | 55+ | -0.04 | 0.07 | 0.56 | 1,067 |
| Sierra Leone 2 | Age | <25 | 55+ | 0.07 | 0.05 | 0.12 | 2,084 |
| Uganda 2 | Age | <25 | 55+ | -0.03 | 0.04 | 0.51 | 1,363 |
| USA | Age | <25 | 55+ | 0.18 | 0.04 | 0.00 | 1,956 |
| Burkina Faso | Education | Secondary + | Up to secondary | 0.09 | 0.03 | 0.00 | 975 |
| Colombia | Education | Secondary + | Up to secondary | -0.05 | 0.03 | 0.10 | 1,009 |
| India | Education | Secondary + | Up to secondary | -0.02 | 0.04 | 0.60 | 338 |
| Mozambique | Education | Secondary + | Up to secondary | 0.04 | 0.03 | 0.24 | 826 |
| Pakistan 1 | Education | Secondary + | Up to secondary | -0.10 | 0.03 | 0.00 | 1,619 |
| Pakistan 2 | Education | Secondary + | Up to secondary | -0.07 | 0.03 | 0.00 | 1,490 |
| Russia | Education | Secondary + | Up to secondary | -0.01 | 0.01 | 0.31 | 22,123 |
| Rwanda | Education | Secondary + | Up to secondary | 0.16 | 0.03 | 0.00 | 1,353 |
| Sierra Leone 1 | Education | Secondary + | Up to secondary | 0.06 | 0.03 | 0.03 | 1,036 |
| Sierra Leone 2 | Education | Secondary + | Up to secondary | -0.01 | 0.02 | 0.64 | 2,108 |
| Uganda 1 | Education | Secondary + | Up to secondary | 0.05 | 0.03 | 0.07 | 2,737 |
| Uganda 2 | Education | Secondary + | Up to secondary | 0.11 | 0.03 | 0.00 | 1,364 |
| USA | Education | Secondary + | Up to secondary | -0.21 | 0.03 | 0.00 | 1,957 |
| Burkina Faso | Gender | Female | Male | 0.06 | 0.03 | 0.06 | 975 |
| Colombia | Gender | Female | Male | 0.04 | 0.03 | 0.18 | 1,010 |
| India | Gender | Female | Male | 0.02 | 0.02 | 0.22 | 1,678 |
| Mozambique | Gender | Female | Male | 0.05 | 0.02 | 0.02 | 860 |
| Nepal | Gender | Female | Male | 0.00 | 0.01 | 0.98 | 1,322 |
| Nigeria | Gender | Female | Male | 0.02 | 0.02 | 0.30 | 1,866 |
| Pakistan 1 | Gender | Female | Male | 0.08 | 0.03 | 0.01 | 1,627 |
| Russia | Gender | Female | Male | 0.16 | 0.01 | 0.00 | 22,123 |
| Rwanda | Gender | Female | Male | 0.09 | 0.02 | 0.00 | 1,353 |
| Sierra Leone 1 | Gender | Female | Male | 0.06 | 0.03 | 0.03 | 1,068 |
| Sierra Leone 2 | Gender | Female | Male | -0.01 | 0.02 | 0.59 | 2,108 |
| Uganda 2 | Gender | Female | Male | 0.03 | 0.02 | 0.18 | 1,364 |
| USA | Gender | Female | Male | 0.17 | 0.03 | 0.00 | 1,957 |

Table 11 shows differences of means between groups within single studies. Estimates are calculated through OLS and represent the difference in the average acceptance rate between the subgroup in column Group and that in column Baseline category.

## Appendix B: Question wording and answer options per study

**Table 12:**
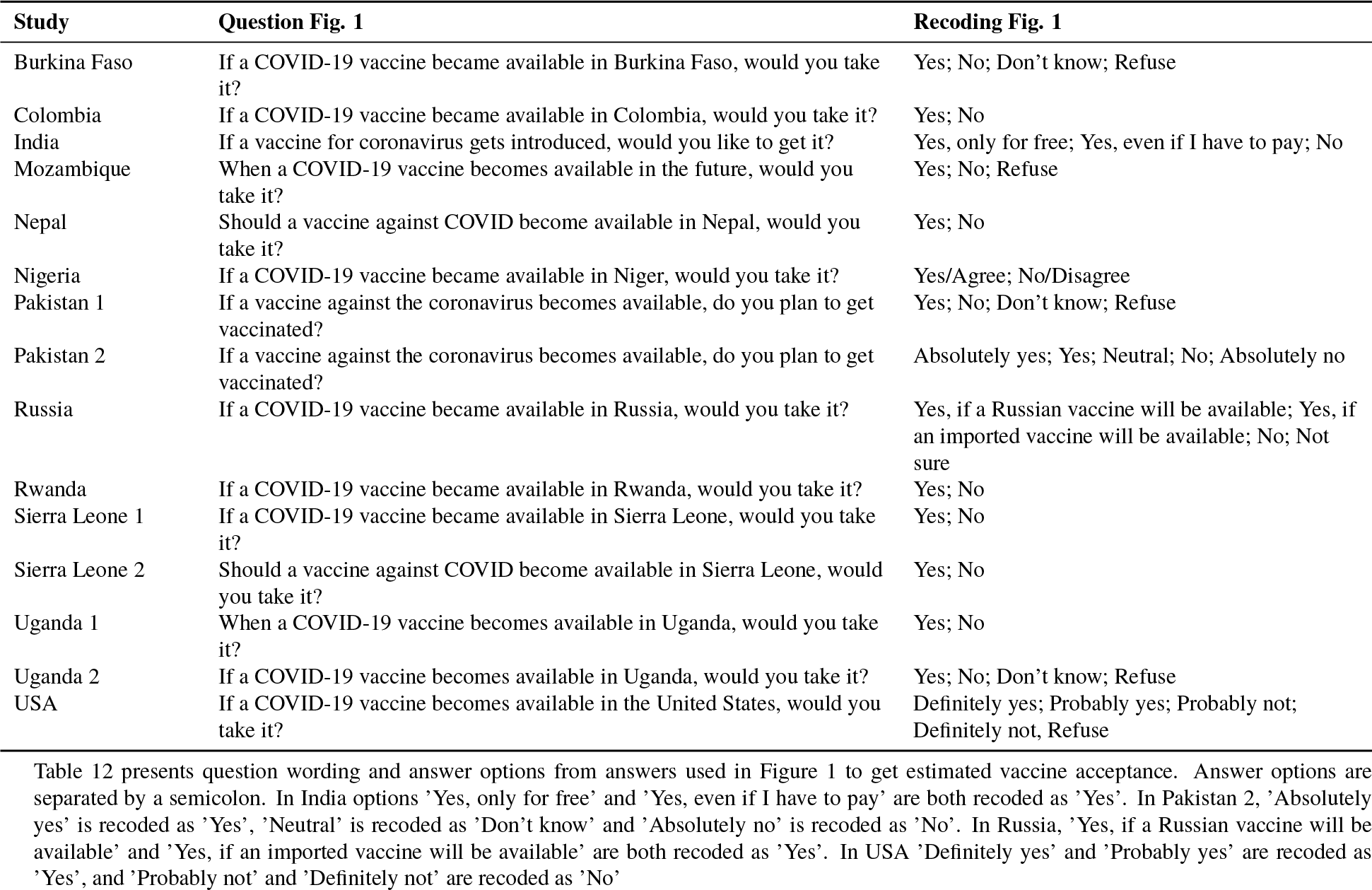
Question wording and answer options: vaccine acceptance.

**Table 13:**
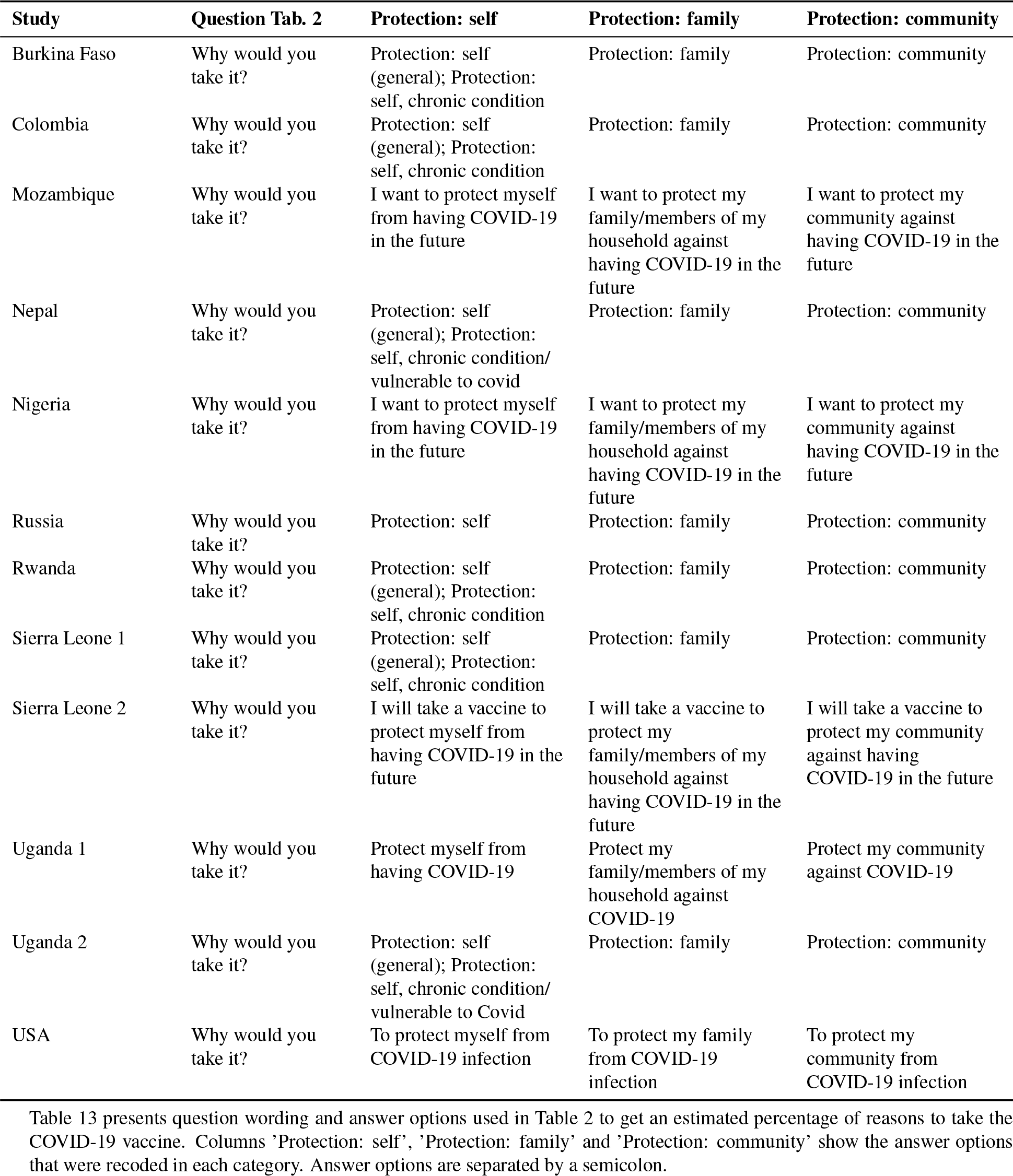
Question wording and answer options: reasons to take vaccine.

**Table 14:**
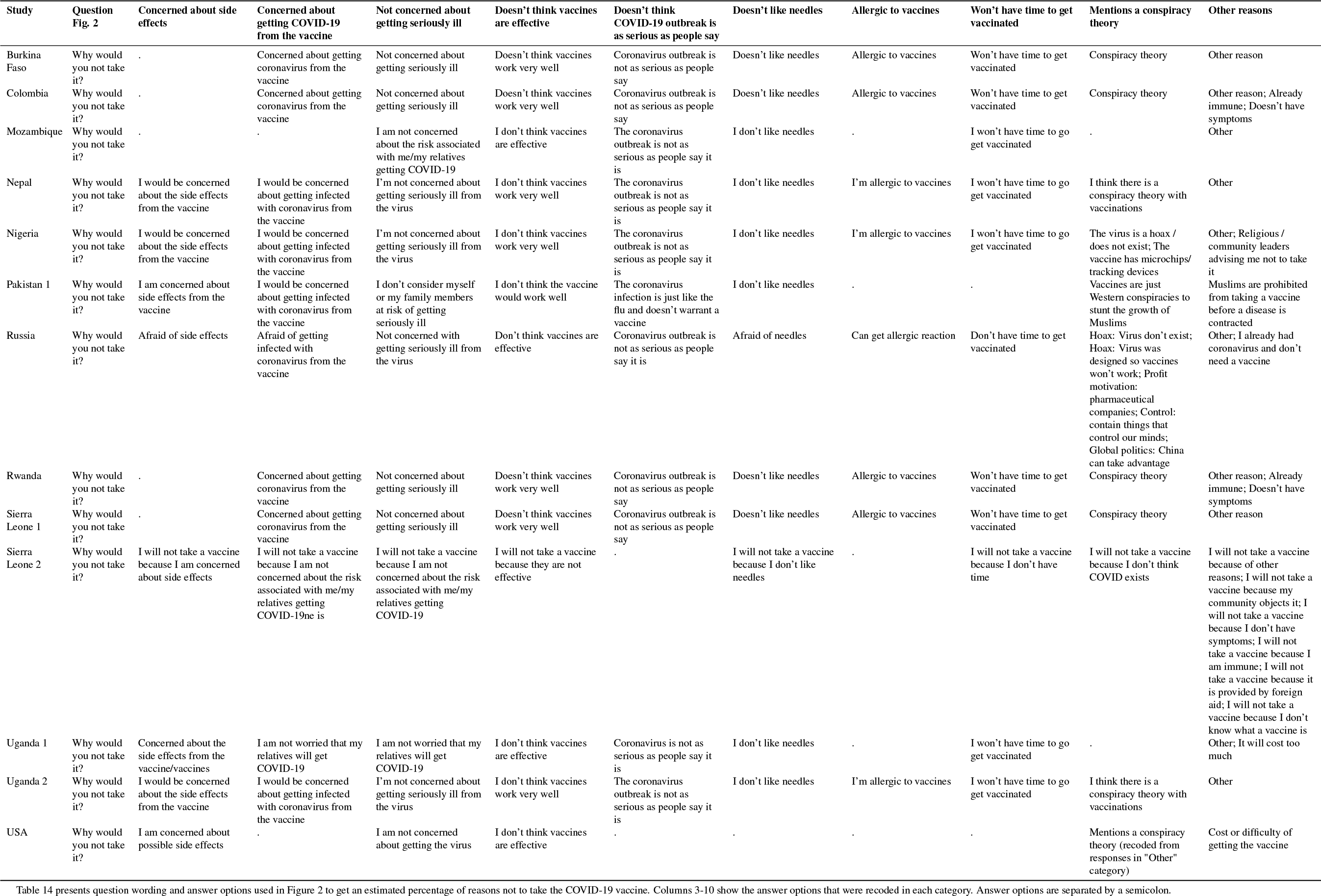
Question wording and answer options: reasons not to take the vaccine.

**Table 15:**
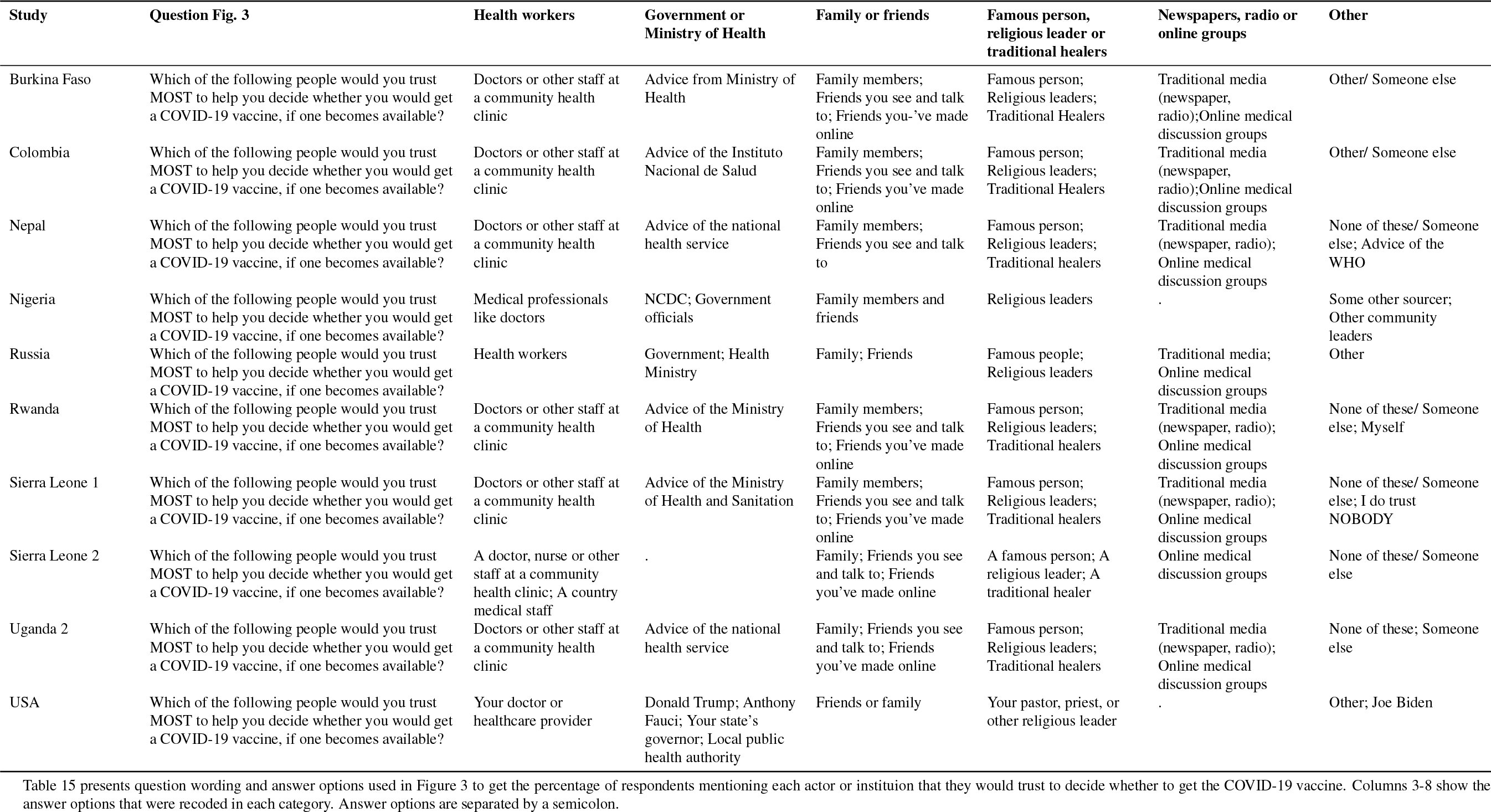
Question wording and answer options: trusted actors and institutions.

| Study | Question Fig. 3 | Health workers | Government or Ministry of Health | Family or friends | Famous person, religious leader or traditional healers | Newspapers, radio or online groups | Other |
| --- | --- | --- | --- | --- | --- | --- | --- |
| Burkina Faso | Which of the following people would you trust MOST to help you decide whether you would get a COVID-19 vaccine, if one becomes available? | Doctors or other staff at a community health clinic | Advice from Ministry of Health | Family members; Friends you see and talk to; Friends you've made online | Famous person; Religious leaders; Traditional Healers | Traditional media (newspaper, radio); Online medical discussion groups | Other/ Someone else |
| Colombia | Which of the following people would you trust MOST to help you decide whether you would get a COVID-19 vaccine, if one becomes available? | Doctors or other staff at a community health clinic | Advice of the Instituto Nacional de Salud | Family members; Friends you see and talk to; Friends you've made online | Famous person; Religious leaders; Traditional Healers | Traditional media (newspaper, radio); Online medical discussion groups | Other/ Someone else |
| Nepal | Which of the following people would you trust MOST to help you decide whether you would get a COVID-19 vaccine, if one becomes available? | Doctors or other staff at a community health clinic | Advice of the national health service | Family members; Friends you see and talk to | Famous person; Religious leaders; Traditional healers | Traditional media (newspaper, radio); Online medical discussion groups | None of these/ Someone else; Advice of the WHO |
| Nigeria | Which of the following people would you trust MOST to help you decide whether you would get a COVID-19 vaccine, if one becomes available? | Medical professionals like doctors | NCDC; Government officials | Family members and friends | Religious leaders | . | Some other sourcer; Other community leaders |
| Russia | Which of the following people would you trust MOST to help you decide whether you would get a COVID-19 vaccine, if one becomes available? | Health workers | Government; Health Ministry | Family; Friends | Famous people; Religious leaders | Traditional media; Online medical discussion groups | Other |
| Rwanda | Which of the following people would you trust MOST to help you decide whether you would get a COVID-19 vaccine, if one becomes available? | Doctors or other staff at a community health clinic | Advice of the Ministry of Health | Family members; Friends you see and talk to; Friends you've made online | Famous person; Religious leaders; Traditional healers | Traditional media (newspaper, radio); Online medical discussion groups | None of these/ Someone else; Myself |
| Sierra Leone 1 | Which of the following people would you trust MOST to help you decide whether you would get a COVID-19 vaccine, if one becomes available? | Doctors or other staff at a community health clinic | Advice of the Ministry of Health and Sanitation | Family members; Friends you see and talk to; Friends you've made online | Famous person; Religious leaders; Traditional healers | Traditional media (newspaper, radio); Online medical discussion groups | None of these/ Someone else; I do trust NOBODY |
| Sierra Leone 2 | Which of the following people would you trust MOST to help you decide whether you would get a COVID-19 vaccine, if one becomes available? | A doctor, nurse or other staff at a community health clinic; A country medical staff | . | Family; Friends you see and talk to; Friends you've made online | A famous person; A religious leader; A traditional healer | Online medical discussion groups | None of these/ Someone else |
| Uganda 2 | Which of the following people would you trust MOST to help you decide whether you would get a COVID-19 vaccine, if one becomes available? | Doctors or other staff at a community health clinic | Advice of the national health service | Family; Friends you see and talk to; Friends you've made online | Famous person; Religious leaders; Traditional healers | Traditional media (newspaper, radio); Online medical discussion groups | None of these; Someone else |
| USA | Which of the following people would you trust MOST to help you decide whether you would get a COVID-19 vaccine, if one becomes available? | Your doctor or healthcare provider | Donald Trump; Anthony Fauci; Your state's governor; Local public health authority | Friends or family | Your pastor, priest, or other religious leader | . | Other; Joe Biden |

## Appendix C: Sample descriptions

The case history data for all countries in our sample is extracted from the Johns Hopkins University Center for Systems Science and Engineering (JHU CSSE) database.^1^

### Burkina Faso, Research for Effective COVID-19 Responses (RECOVR) National RDD Sample, Innovations for Poverty Action (IPA)

#### COVID-19 Experience

- First confirmed case: March 9, 2020
- Number of confirmed cases 2,335 as of October 15, 2020
- Number of deaths: 65 as of October 15, 2020

#### Target Population

A random sample of all adults with mobile phone numbers in the country, based on national communications authority number allocation plans.

#### Original Study Design

N/A

#### COVID-19 Survey Design

Numbers were called via random digit dialing (RDD), stratified by mobile network operator market share for a two-round panel survey.

*Sampling Frame:* All mobile phone numbers in Burkina Faso.

*Survey Dates:* October 15 to December 4, 2020 (Round 1 June 6-15, 2020)

*Sample size, tracking and attrition:* Sample includes 977 respondents from the second round of a panel. In the first round conducted between June 6 to 15, 2020, 1,356 individual surveys were contacted through Random Digit Dialing (RDD) from the sampling frame of all mobile phone numbers in Burkina Faso. 2,313 working numbers yielded 1,383 eligible respondents for a completion rate of 98% of eligible respondents.

*Sampling Weights:* Post-stratification weights are computed to adjust for differential attrition between the first and second rounds of the RDD panel, weighting on gender, region, and educational attainment.

*IRB Approval:* This research was approved via IPA IRB Protocol 15608, and the Burkina Faso Institutional Ethics Committee for Health Sciences Research, approval A13-2020.

### Colombia, Research for Effective COVID-19 Responses (RECOVR) National RDD Sample, Innovations for Poverty Action (IPA)

#### COVID-19 Experience

- First confirmed case: March 6, 2020
- Number of confirmed cases: 456,689 as of August 15, 2020
- Number of deaths: 14,810 as of August 15, 2020

#### Target Population

A random sample of all numerically possible mobile phone numbers in the country, based on national communications authority number allocation plans.

#### Original Study Design

N/A

#### COVID-19 Survey Design

*Sampling Frame:* Numbers were called via random digit dialing (RDD), stratified by mobile network operator market share.

*Survey Dates:* August 15-25, 2020 (Round 1 May 8-15, 2020)

*Sample size, tracking and attrition:* Sample includes 1,012 respondents contacted in the second round of a panel of 1,507.

*Sampling Weights:* Post-stratification weights are computed to adjust for differential attrition between the first and second rounds of the RDD panel, weighting on gender, region, and educational attainment.

*IRB Approval:* This research was approved via IPA IRB Protocol 15582.

### India, Coping with COVID-19 in Slums: Evidence from India Subnational sample, Nova School of Business and Economics, The Institute for Fiscal Studies, University of St. Andrews

#### COVID-19 Experience

- First confirmed case: January 30, 2020
- Number of confirmed cases: 198,370 as of June 1, 2020
- Number of deaths: 5,608 as of June 1, 2020

#### Target Population

Random subset of slum populations in Lucknow and Kanpur, Uttar Pradesh, India. Socio-economic variables are only collected for a representative sample of the population relying on community toilets or open defecation to fulfil their sanitation needs.

#### Original Study Design

Randomized controlled trial, with complete census of households within 142 slums (September to December 2017), and a series of household and caretaker surveys, objective measurements, incentivized behavioural measurements, and a Structured Community Activity, collected for a sub-set of 100 slums between April 2018 and September 2019.

*Intervention:* Catchment areas of CTs were randomly allocated to two interventions. The first intervention aimed at community toilet improvements by offering caretakers the choice of a grant to be spent for improvements in the facility. Following the grant, caretakers were offered a large financial reward conditional on the cleanliness of the facility. The second intervention added to this CT improvement awareness creation among potential users through face-to-face information sessions, leaflets, monthly reminders using voice messages sent to mobile phones, and posters hung in the CTs.

*Sampling Frame:* A two-step sampling was applied, first, study households from the main study sample were sampled, then households from the whole slum population were added.

*Survey Dates:* Baseline: June to July 2020, Follow-up 1: October to November 2020, Follow-up 2: December 16, 2020 to January 18, 2021.

*Sample size, tracking and attrition:* 3,991 households, with a mean of 28 households per cluster (142). Non-response Baseline: 25%, Attrition rate Baseline to Follow-up (1 and 2): 13%, Randomly selected replacement households for Follow-up (1 and 2): 1,277.

*Sampling Weights:* Included

*IRB Approval:* Approval was secured from London School of Economics (REC ref. 1132). The pre-analysis plan was registered on the AEA RCT registry (RCT ID AEARCTR-0006564).

### Mozambique Subnational sample, International Growth Center, Nova School of Business and Economics

#### COVID-19 Experience

- First confirmed case: March 22, 2020
- Number of confirmed cases: 12,777 as of October 30, 2020
- Number of deaths: 91 as of October 30, 2020

#### Target Population

Microentrepreneurs in urban markets of Maputo and household heads from the province of Cabo Delgado.

#### Original Study Design

Initial data were collected in-person in two different studies. For microentrepreneurs in Maputo, the data were collected between October 2013 and April 2014 (baseline), and between July and November 2015 (endline).^2^ For household heads in Cabo Delgado, the data were collected in-person between August and September 2016 (baseline), and between August and September 2017 (endline).^3^

*Intervention:* The first study was dedicated to analyzing the impacts of interventions targeting microentrepreneurs in urban markets on financial inclusion and literacy. The second study focused on the role of information to counteract the political resource curse after a substantial natural gas discovery.

*Sampling Frame:* The first initial sample was selected by in-field random sampling in 23 urban and periurban markets in Maputo and Matola. Stratification was based on the gender of the respondent and on the type of establishment (stall vs. store). The second initial sample was selected to be representative of 206 communities in the province of Cabo Delgado, randomly drawn from the list of all 421 polling locations in the sampling frame, stratified on urban, semiurban, and rural areas. This survey in this paper was done by phone.

*Survey Dates:* October 30 to November 21, 2020 (Maputo) and November 6 to November 30, 2020 (Pemba).

*Sample size, tracking and attrition:* 554 microentrepreneurs from Maputo and 308 households from Cabo Delgado.

*Sampling Weights:* N/A

\emph{IRB Approval: The approval was secured from Universidade Nova de Lisboa on July 14, 2020.

### Nepal, Western Terai Panel Survey (WTPS) Subnational sample, Yale University, Yale Research Initiative on Innovation and Scale (Y-RISE)

#### COVID-19 Experience

- First confirmed case: January 23, 2020
- Number of confirmed cases: 233,452 as of December 1, 2020
- Number of deaths: 1,529 as of December 1, 2020

#### Target Population

Rural households in the districts of Kailali and Kanchanpur.

#### Original Study Design

Initial baseline data was collected in-person in July of 2019, and 5 rounds of phone survey data were collected between August 12, 2019 and January 4, 2020.

*Sampling Frame*

The phone survey sample includes 2,636 rural households in the districts of Kailali and Kanchanpur, which represent the set of households that responded to phone surveys from an original sample of 2,935 households. This sample was constructed by randomly sampling 33 wards from 15 of the 20 sub-districts in Kailali and Kanchanpur and selecting a random 97 villages from within those wards. At the time of baseline data collection in July of 2019, 7 of these 97 villages were dropped from the sample due to flooding. Households belong to the bottom half of the wealth distribution in these villages, as estimated by a participatory wealth ranking exercise with members of the village.

*Survey Dates:*December 1st - December 11, 2020

*Sample size, tracking and attrition:* 1,392 households

*IRB Approval:* This research was approved via Yale University IRB Protocol 2000025621.

### Nigeria Subnational sample, WZB Berlin Social Science Center, University of Illinois Chicago

#### COVID-19 Experience

- First confirmed case: February 28, 2020
- Number of confirmed cases: 65,693 as of November 18, 2020
- Number of deaths: 1,163 as of November 18, 2020

#### Target Population

Christian and Muslim men and women, age 18 and above, living in Kaduna state, Nigeria.

#### Original Study Design

Initial data was collected from a subset of the sample in December 2019 (in person survey) and July - Aug 2020 (phone survey) as part of an experiment testing the effects of a brief radio program on inter-religious animus. A random walk procedure and random sampling were used within households to recruit a representative sample of adults in Kaduna town. The rest of the sample was recruited for the study in Aug 2020 by purchasing phone lists for residents of Kaduna State.

*Intervention:* The study examines the effects of a radio program and a TV drama on inter-religious animus. The subset of the sample in the radio study was randomly assigned to listen to a brief radio program on one of the following topics: (1) an inter-religious storyline, (2) an intra-religious storyline, and (3) a message about maintaining safe health practices. All respondents in the sample participated in a study examining the effect of viewing an inter-religious storyline unfolding over a full season of a popular TV drama, Dadin Kowa. The season aired from Aug - Oct 2020. A third of the sample were encouraged to watch Dadin Kowa, a third were encouraged to watch the TV station Africa Magic Hausa at the same time Dadin Kowa aired, and a third were in the treatment-as-usual group. All participants received a weekly incentivized SMS quiz from Aug - Oct 2020.

#### COVID-19 Survey Design

This survey is not primarily about COVID-19, but was designed as an endline survey to follow the TV drama intervention described above. The goal of this survey is to measure a range of attitudinal outcomes related to Christian-Muslim relations (including prejudice, intergroup threat perceptions, dehumanization, and support for the use of violence, among others). We included nine of the standardized COVID-19 vaccine-related questions collected specifically for this vaccine acceptance study in the final module of the endline survey.

*Sampling Frame:* 950 respondents in the sample were recruited in person through a random sampling procedure in the Kaduna metropolitan area (pre-COVID). The remaining 1,700 respondents were recruited into the study over the phone from lists of phone numbers of Kaduna state residents that were purchased from a private vendor.

*Survey Dates:* November 18 - December 18, 2020.

*Sample size, tracking and attrition:* All 1,834 individuals who completed the endline survey are included.

*Sampling Weights:* N/A

*IRB Approval:* This study was reviewed by the IRB at the University of Pennsylvania (Protocol 834548), and it was determined on November 20, 2019 to meet the criteria for review exemption (45 CFR 46.104, category #2).

### Pakistan

#### COVID-19 Experience

- March 6: First confirmed case: February 26, 2020
- Number of confirmed cases: 271,887 as of July 24, 2020
- Number of deaths: 5,787 as of July 24, 2020

**Pakistan, Economic Vulnerability Assessment (EVA) Subnational sample, Sheikhupura Police Study Sample, Institute of Development and Economic Alternatives, Lahore University of Management Science, London School of Economics, Princeton University (Pakistan 1)**

#### Target Population

A representative sample of adults from 108 of 151 police beats in Sheikhupura and Nankana districts of Punjab Province.

#### Original Study Design

N/A

#### COVID-19 Survey Design

The EVA survey involved calls to all households in the stratified random sample for the policing study midline survey.

*Sampling Frame:* Households in Sheikhupura and Nankana districts.

*Survey Dates:* July 24 to September 9, 2020

*Sample size, tracking and attrition:* Sample includes 1,473 respondents.

*Sampling Weights:* Post-stratification weights are computed to adjust for the sampling process, which involved stratifying first on 27 police stations, then within each police station on beats, then PPS sampling within beats using Asiapop population data.

*IRB Approval:* This research was approved via Princeton University IRB Protocol 7250.

### Pakistan, Economic Vulnerability Assessment (EVA) Subnational sample (Pakistan 2)

#### Target Population

All possible mobile phone numbers (in the province of Punjab) generated based on the local mobile phone number structure in Pakistan.

#### Original Study Design

N/A

#### COVID-19 Survey Design

The EVA survey involved making calls to individuals in Punjab based on random digit dialing.

*Sampling Frame:* Individuals with mobile phones in Punjab.

*Survey Dates:* September 2 to October 13, 2020

*Sample size, tracking and attrition:* Sample includes 1,492 respondents.

*Sampling Weights:* N/A.

*IRB Approval:* This research was approved by Lahore University of Management Sciences IRB Protocol LUMS-IRB/07012020SA.

### Rwanda, Research for Effective COVID-19 Responses (RECOVR) National RDD Sample, Innovations for Poverty Action (IPA)

#### COVID-19 Experience

- First confirmed case: March 14, 2020
- Total cases: 5,017 as of October 22, 2020
- Total deaths: 34 as of October 22, 2020

**Target Population**: A random sample of all numerically possible mobile phone numbers in the country, based on national communications authority number allocation plans.

#### Original Study Design

N/A

#### COVID-19 Survey Design

Phone survey

*Sampling Frame:* Numbers were called via random digit dialing (RDD), stratified by mobile network operator market share.

*Survey Dates:* October 22 to November 5, 2020 (Round 1 June 4 -12, 2020)

*Sample size, tracking and attrition:*Sample includes 1,355 respondents contacted in the second round of a panel of 1,480.

*Sampling Weights:* Post-stratification weights are computed to adjust for differential attrition between the first and second rounds of the RDD panel, weighting on gender, region, and educational attainment.

*IRB Approval:* This research was approved via IPA IRB Protocol 15591, Rwanda National Institute for Scientific Research permit No.0856/2020/10/NISR; and Rwanda National Ethics Committee approval No.16/RNEC/2020.

### Russian Federation, Research on COVID-19 in Russia’s Regions (RoCiRR) Subnational sample, International Center for the Study of Institutions and Development (HSE University, Moscow, Russia) and Economics Department of Ghent University, WZB Berlin Social Science Center, Columbia University

#### COVID-19 Experience

- First confirmed case: January 31, 2020
- Number of confirmed cases: 1,720,063 as of November 6, 2020
- Number of deaths: 29,654 as of November 6, 2020

#### Target Population

Adult internet users who reside in one of 61 federal subjects (federal cities, oblasts, republics, krais and autonomous okrug) of Russia. The regions included in the study are Republics: *Bashkortostan, Karelia, Komi, Mariy El, Mordovia, Tatarstan, Udmurtia, Chuvashia*. Krais: *Altai, Krasnodarsky, Krasnoyarsky, Permsky, Primorsky, Stavropolsky, Khabarovsky*. Oblasts: *Arkhangelsk, Astrakhan, Belgorod, Bryansk, Vladimir, Volgograd, Vologda, Voronezh, Ivanovo, Irkutsk, Kaliningrad, Kaluga, Kemerovo, Kirov, Kostroma, Kurgan, Kursk, Leningrad, Lipetsk, Moscow, Murmansk, Nizhny Novgorod, Novgorod, Novosibirsk, Omsk, Orenburg, Orel, Pskov, Penza, Rostov, Ryazan, Samara, Saratov, Sverdlovsk, Smolensk, Tambov, Tver, Tomsk, Tula, Tyumen, Ulyanovsk, Chelyabinsk, Yaroslavl*. Other: *Moscow, Saint Petersburg, Khanty-Mansiysk Autonomous Okrug – Ugra*. The remaining 24 federal subjects were excluded from the study due to inability to enroll sample size with desired characteristics (sample size, age, gender and education group composition) and account for less than 14% of the total adult population of Russia.

#### Original Study Design

N/A

#### COVID-19 Survey Design

The study was designed to measure the impact of pandemics on Russians, mostly those who live in cities with more than 100,000 residents. It contains a number of questions on the personal experience, norms and values, trust in government institutions, provision of social services, and mass media use. Region and geolocality of every respondent are recorded.

*Sampling Frame:* In total 25,558 respondents received the module on vaccine acceptance. The sample was enrolled from the pool of Russian online survey company OMI (Online Market Intelligence). The sampling was specifically targeted at having a minimum of 150 respondents in each of the 61 regions and including respondents from all the main age and gender groups within each region. Respondents were also selected so that at least 40% of respondents did not have higher education, in accordance with higher education rates in Russia. Out of 25,558 recruited respondents, 22,125 completed the survey. Among 22,125 respondents who completed the survey, 20,821 were enrolled from the general pull of the survey company respondents, while the remaining 1,304 respondents were enrolled among residents of cities with populations below 100,000 and rural areas.

*Survey Dates:* November 6 - December 1, 2020

*Sample size, tracking and attrition:* 22,125 respondents who completed the survey with the vaccine acceptance module included.

*Sampling Weights:* Post-stratification weights are computed to match marginal distributions of age, gender and education among the adult population of Russia with target proportions coming from the 2019 Yearbook and 2015 Microcensus released by Russian Federal Bureau of National Statistics (Rosstat).

*IRB Approval:* This study was approved via Columbia University IRB Protocol IRB-AAAT4453.

### Sierra Leone

#### COVID-19 Experience

- First confirmed case: March 20, 2020
- Total cases: 2,252 as of October 2, 2020 and 3,030 as of January 20, 2021
- Total deaths: 72 as of October 2, 2020 and 77 as of January 20, 2021

**Sierra Leone, Research for Effective COVID-19 Responses (RECOVR) National RDD Sample, Innovations for Poverty Action (IPA) (Sierra Leone 1) Target Population**: A random sample of all numerically possible mobile phone numbers in the country, based on national communications authority number allocation plans.

#### Original Study Design

N/A

#### COVID-19 Survey Design

Numbers were called via random digit dialing (RDD), stratified by mobile network operator market share

*Sampling Frame:* All active mobile phone numbers in Sierra Leone.

*Survey Dates:* October 2-19, 2020 (Round 1 May 27 to June 15, 2020)

*Sample Size, tracking and Attrition:* Sample includes 1,070 respondents contacted in the second round of a panel of 1,304.

*Sampling Weights:* Post-stratification weights are computed to adjust for differential attrition between the first and second rounds of the RDD panel, weighting on gender, region, and educational attainment.

*IRB Approval:* This research was approved via IPA IRB Protocol 15592, and Sierra Leone Ethics and Scientific Review Committee approval (no approval number, letter available upon request).

### Sierra Leone, Towns that are Candidates for Rural Electrification Nation-wide sample, International Growth Centre (IGC), Wageningen University & Research, Yale Research Initiative on Innovation and Scale (Y-RISE), WZB Berlin Social Science Center and Columbia University (Sierra Leone 2) Project Title: Sierra Leone Rural Electrification (SLRE)

#### Target Population

Households in 195 rural towns across all 14 districts of Sierra Leone. Of these, 97 villages were selected to benefit from an electrification program.

#### Original Study Design

Initial baseline data was collected during late 2019 and early 2020 as part of a study to assess the impact of Rural Electrification in rural towns in Sierra Leone.

*Intervention:* The Government of Sierra Leone (GoSL) in collaboration with the United Nations Office for Project Services (UNOPS) and international donors is implementing the Rural Renewable Energy Project (RREP). In its first wave, during 2017, the project provided stand-alone solar photovoltaic powered mini-grids to 54 communities across the country. Construction of mini-grids in a further 43 towns is ongoing. In RREP communities, engineers construct 6kW–36kW power mini-grids that provide reliable power year-round. Electricity is free for schools and clinics. Residential and commercial users can acquire connections from commercial operators.

*Village Sampling Frame:* Household data was collected in 195 towns across all 12 districts of Sierra Leone. The GoSL selected 97 towns with (planned) mini-grids. We used Propensity Score Matching to select 98 control communities. Within communities, respondents were randomly selected from a census roster stratified by occupation status of farmers, business owners and other occupations [47 percent, 47 percent and 7 percent]. In each village, the intended sample was 43 households (20 farmers, 20 businesses, 3 others). Data was collected during June–July (108 communities) and November–December 2019 (87 communities). If a household on the sampling list was not available on the village visit day, we had a randomly sampled list of replacement households to survey. The replacement household would be the same occupation as the sampled household would have been so the sample ratio of 20-20-3 still held in each community.

#### COVID-19 Survey Design

The goal was to assess households’ degree of economic vulnerability in the face of the COVID-19 pandemic.

*Sampling Frame:* The COVID-19 survey data comprises 2,110 respondents from 186 towns from the original baseline survey. Phone surveys were attempted to all 195 rural communities from the baseline survey. The total baseline household sample comprised 7047 respondents. We recontacted all baseline respondents that listed a phone number (4,594 respondents) and obtained informed consent for the phone survey. We implemented several waves of the phone survey, recontracting a respondent about every month. In wave 7, we added questions related to Vaccine Acceptability.^4^.

*Survey Dates:* October 7, 2020 to January 20, 2021 (earlier rounds included Wave 1: April 29-May

15; Wave 2: May 15-June 4; Wave 3: June 5-June 17; Wave 4: June 17-June 30; Wave 5: July 1-August 8; Wave 6: August 19-October 1). The median survey time was 33 minutes.

*Sample size, tracking and attrition:* Data collection took place between October 7 and January 20, 2021with 2,110 respondents, in 186 towns for a tracking rate of 46 percent.

*Sampling Weights:* None

*IRB Approval:* Approval was secured from Sierra Leone Ethics and Scientific Review Committee (SLERC 2904202) and Wageningen University (24062020).

### Uganda

#### COVID-19 Experience

- First confirmed case: March 21, 2020
- Total cases: 741 as of June 18, 2020 and 6,468 as of September 21, 2020
- Total deaths: 0 as of June 18, 2020 and 63 as of September 21, 2020

**Uganda Subnational sample, International Growth Center, Trinity College Dublin, Stockholm School of Economics and Misum, Institute for International Economic Studies, Stockholm University (Uganda 1) Target Population**: Women from semi-rural and rural villages across 13 districts in Uganda (Iganga, Kayunga, Mbale, Mityana, Apac, Dokolo, Gulu, Adjumani, Koboko, Maracha, Nebbi, Soroti, Kumi).

#### Original Study Design

Initial baseline data was collected in 2016 as part of a large cluster randomized controlled trial, with the aim of selecting households likely to have children during the study period. Four criterias for selection were thus used, in descending order of importance: the household has a woman that is currently pregnant, or aged 16-30 years old, with a young child less than three years old, and/or married (formally or informally). In each household, the respondent was chosen as the female household head or the primary female health care giver of the household if the household head could not be found.

#### COVID-19 Survey Design

The data was collected through multiple rounds of phone surveys. The variable measuring age was constructed by approximation, using the baseline data from 2016 and adding 4 years to the 2016 measure. When the baseline respondent was replaced, the initial age information was deleted.

*Sampling Frame:* Households were selected within 500 clusters (the village of the household).

*Survey Dates:* September 21 to December 06, 2020.

*Sample size, tracking and attrition:* Out of 2,743 respondents, 1752 were included, provided that they answered the main question about vaccine uptake.

*Sampling Weights:* None.

*IRB Approval:* Mildmay Uganda Research Ethics Committee (protocol number 0109-2015) on September 21, 2020.

### Uganda Subnational sample, WZB Berlin Social Science Center and Columbia University, NYU Abu Dhabi, Innovations for Poverty Action (IPA) (Uganda 2)

#### Target Population

All residents of Kampala who are Ugandan citizens, above the age of 18, and agree in principle to attend a short citizen consultative meeting.

Original Study Design

Baseline data was collected between July and October 2019 for an intervention that randomized citizen attendance to a set of 188 consultative meetings organized across Kampala. The meetings were organized to collect citizen preferences for the design of a forthcoming municipal citizen charter. The study also aimed to assess patterns of political inequality in meeting participation, dynamics, and outcomes, as well as study the subsequent effects on prosociality of being incorporated in this participatory process. 1/3 of the sample was randomly allocated to control, while 2/3 of respondents were invited to attend a consultative meeting. The consultations took place between November 2019 and February 2020 across Kampala divisions.

*Intervention:* The intervention consisted of attendance at the consultative meeting organized a few months after baseline data collection. A further randomization allocated ½ of the invited participants to a meeting moderated by a local bureaucrat, while the remaining ones attended a meeting moderated by a neutral discussion leader.

COVID-19 Survey Design: The COVID-19 survey sample comprises the 2,189 respondents to the baseline who were selected on the basis of their residence in the city. Having received permission to re-contact these individuals, we coordinated a 3-wave panel throughout the summer and fall of 2020, with respondents contacted via phone. The goal was to assess households’ degree of economic vulnerability in the face of the COVID-19 pandemic and respondents’ evaluations of performance of political actors in tackling the pandemic.

*Sampling Frame:* The 2,189 respondents to the baseline were randomly selected from a sampling frame of all buildings in Kampala, for which information about their geographical coordinates was available. After randomly selecting a set of candidate structures, interviewers sampled respondents from the subset of structures that were residential.

*Survey Dates:* Wave 1: June 18–July 23. Wave 2: September 4–29. Wave 3: November 23–December 12.

*Sample size, tracking and attrition:* Of the 2,189 respondents which we aimed to contact, we were able to reach 1,333 in Wave 1, 1,289 in Wave 2, and 1,366 in Wave 3. Wave 3 contained the COVID-19 vaccine module presented in this analysis.

*Sampling Weights:* None.

*IRB Approval:* The study was approved by IPA Global IRB (protocol number 15018) on May 29, 2020; WZB Berlin Social Science Center Ethics Review Board (protocol number 2020/0/91) on June 10, 2020; NYU Abu Dhabi IRB (protocol number HRPP-2020-64) on May 27, 2020; MIT Committee on the Use of Humans as Experimental Subjects (protocol number 2005000155) on June 3, 2020; and by the Mildmay Uganda Research Ethics Committee (protocol number 0604-2019) on June 11, 2020.

### United States of America Nation-wide sample, WZB Berlin Social Science Center, Cornell University, Tufts University

#### COVID-19 Experience

- First confirmed case: January 20, 2020
- Total cases: 14,499,637 as of December 4, 2020
- Total deaths: 281,678 as of December 4, 2020

#### Target Population

Nation-wide sample of adult internet users recruited through the market research firm Lucid.

#### Original Study Design

N/A

*Intervention:* N/A

#### COVID-19 Survey Design

This survey was part of a panel study on attitudes toward COVID-19 technologies and public health surveillance.

*Sampling Frame:* The Lucid Marketplace is an automated marketplace that connects researchers with willing online research participants. Lucid partners with a network of companies that maintain relationships with research participants by engaging them with research opportunities. While Lucid does not provide probability samples of the U.S. adult population, its quota samples approximate the marginal distributions of key demographic characteristics. Recent validation exercises have found that Lucid samples approximate nationally representative samples in terms of demographic characteristics and survey experiment effects.^5^

*Survey Dates:* December 4-5, 2020

*Sample size, tracking and attrition:* 1,959 individual online surveys. In the main question regarding intention to take the vaccine, approximately 10% of respondents (184) did not answer *Sampling Weights:* Post-stratification weights are computed to match marginal population distributions of income, age, education, gender, race and region among the US adult population, with target proportions based on the 2018 American Community Survey.

*IRB Approval:* This study received approval from the Cornell University IRB under Protocol #2004009569.

## Footnotes

1 Dong, E., Du, H., & Gardner, L. (2020). An interactive web-based dashboard to track COVID-19 in real time. The Lancet infectious diseases, 20(5), 533-534.

2 Original study: http://catiabatista.org/bsv_mm_urban.pdf

3 Original study: https://www.aeaweb.org/articles?id=10.1257/aer.20190842

4 The data was first reported in https://www.theigc.org/wp-content/uploads/2020/05/Meriggi-et-al-Data-Brief-2020.pdf

5 See for instance: https://journals.sagepub.com/doi/10.1177/2053168018822174

## References

1. World Health Organization. Draft landscape and tracker of COVID-19 candidate vaccines. https://www.who.int/publications/m/item/draft-landscape-of-covid-19-candidate-vaccines (2021).

2. McGill COVID19 Vaccine Tracker Team. COVID-19 Vaccine Tracker. https://covid19.trackvaccines.org/vaccines/ (2021).

3. Wouters, O. J., et al. Challenges in ensuring global access to COVID-19 vaccines: Production, affordability, allocation, and deployment. The Lancet (2021).

4. de Figueiredo, A., Simas, C., Karafillakis, E., Paterson, P. & Larson, H. J. Mapping global trends in vaccine confidence and investigating barriers to vaccine uptake: A large-scale retrospective temporal modelling study. The Lancet 396, 898–908 (2020).

5. Ipsos-World Economic Forum. Global attitudes: COVID-19 vaccines. https://www.ipsos.com/en-ro/global-attitudes-covid-19-vaccine-january-2021 (2021).

6. Malik, A., McFadden, S., Elharake, J. & Omer, S. Determinants of COVID-19 vaccine acceptance in the US. EClinicalMedicine 26, 100495 (2020).

7. Kreps, S. et al. Factors associated with US adults’ likelihood of accepting COVID-19 vaccination. JAMA Network Open 3, e2025594–e2025594 (2020).

8. Shekhar, R. et al. COVID-19 vaccine acceptance among health care workers in the United States. Vaccines 9, 119 (2021).

9. Dror, A. A. et al. Vaccine hesitancy: the next challenge in the fight against COVID-19. European Journal of Epidemiology 35, 775–779 (2020).

10. Fisher, K. A. et al. Attitudes toward a potential SARS-CoV-2 vaccine: a survey of US adults. Annals of Internal Medicine 173, 964–973 (2020).

11. Lazarus, J. V. et al. A global survey of potential acceptance of a COVID-19 vaccine. Nature medicine 27, 225–228 (2021).

12. Ong, S. W. X., Young, B. E. & Lye, D. C. Lack of detail in population-level data impedes analysis of SARS-CoV-2 variants of concern and clinical outcomes. The Lancet Infectious Diseases (2021).

13. The Lancet COVID-19 Comission Task Force on Public Health Measures to Suppress the Pandemic. SARS-CoV-2 variants: the need for urgent public health action beyond vaccines. https://covid19commission.org/public-health-measures (2021).

14. Mukherjee, S. Why does the pandemic seem to be hitting some countries harder than others? New Yorker (2021).

15. Brewer, N. T. et al. Meta-analysis of the relationship between risk perception and health behavior: The example of vaccination. Health psychology 26, 136 (2007).

16. Christensen, D., Dube, O., Haushofer, J., Siddiqi, B. & Voors, M. Building Resilient Health Systems: Experimental Evidence from Sierra Leone and the 2014 Ebola Outbreak. The Quarterly Journal of Economics 136, 1145–1198 (2021).

17. Lowes, S. & Montero, E. The legacy of colonial medicine in Central Africa. American Economic Review 111, 1284–1314 (2021).

18. Martinez-Bravo, M. & Stegmann, A. In Vaccines We Trust? The Effects of the Cia’s Vaccine Ruse on Immunization in Pakistan. CEPR Discussion Paper No. DP15847 (2021).

19. Jegede, A. S. What led to the Nigerian boycott of the polio vaccination campaign? PLOS Medicine 4(3), (2007).

20. Blair, R., Morse, B. & Tsai, L. Public health and public trust: Survey evidence from the Ebola Virus Disease epidemic in Liberia. Social Science & Medicine 172, 89–97 (2017).

21. Ali, M. et al. Polio vaccination controversy in pakistan. The Lancet 394, 915–916 (2019).

22. Robbins, A. The CIA’s vaccination ruse. Journal of Public Health Policy 33, 387–389 (2012).

23. Deserranno, E. & León-Ciliotta, G. Promotions and Productivity: The Role of Meritocracy and Pay Progression in the Public Sector. CEPR Discussion Paper No. DP15837 (2021).

24. Betsch, C. et al. Beyond confidence: Development of a measure assessing the 5C psychological antecedents of vaccination. PloS one 13, e0208601 (2018).

25. Domek, G. J. et al. Measuring vaccine hesitancy: Field testing the WHO SAGE working group on vaccine hesitancy survey tool in guatemala. Vaccine 36, 5273–5281 (2018).

26. Shapiro, G. K. et al. The vaccine hesitancy scale: Psychometric properties and validation. Vaccine 36, 660–667 (2018).

27. Gilkey, M. B. et al. The vaccination confidence scale: A brief measure of parents’ vaccination beliefs. Vaccine 32, 6259–6265 (2014).

28. World Health Organization. Data for action: achieving high uptake of COVID-19 vaccines. https://www.who.int/publications/i/item/WHO-2019-nCoV-vaccination-demand-planning-2021.1 (2021).

29. Chou, W.-Y. S. & Budenz, A. Considering emotion in COVID-19 vaccine communication: Addressing vaccine hesitancy and fostering vaccine confidence. Health communication 35, 1718–1722 (2020).

30. Mulligan, M. J. et al. Phase i/II study of COVID-19 RNA vaccine BNT162b1 in adults. Nature 586, 589–593 (2020).

31. Logunov, D. Y. et al. Safety and efficacy of an rAd26 and rAd5 vector-based heterologous prime-boost COVID-19 vaccine: An interim analysis of a randomised controlled phase 3 trial in russia. The Lancet 397, 671–681 (2021).

32. Folegatti, P. M. et al. Safety and immunogenicity of the ChAdOx1 nCoV-19 vaccine against SARS-CoV-2: A preliminary report of a phase 1/2, single-blind, randomised controlled trial. The Lancet 396, 467–478 (2020).

33. Voysey, M. et al. Safety and efficacy of the ChAdOx1 nCoV-19 vaccine (AZD1222) against SARS-CoV-2: An interim analysis of four randomised controlled trials in brazil, south africa, and the UK. The Lancet 397, 99–111 (2021).

34. Baden, L. R. et al. Efficacy and safety of the mRNA-1273 SARS-CoV-2 vaccine. New England Journal of Medicine 384, 403–416 (2021).

35. Polack, F. P. et al. Safety and efficacy of the BNT162b2 mRNA covid-19 vaccine. New England Journal of Medicine 383, 2603–2615 (2020).

36. Wadman, M. Public needs to prep for vaccine side effects. Science 370, 1022–1022 (2020).

37. Callaway, E. Russia’s fast-track coronavirus vaccine draws outrage over safety. Nature 584, 334–335 (2020).

38. Stein, R. A. The golden age of anti-vaccine conspiracies. Germs 7, 168 (2017).

39. Loomba, S., de Figueiredo, A., Piatek, S. J., Graaf, K. de & Larson, H. J. Measuring the impact of COVID-19 vaccine misinformation on vaccination intent in the UK and USA. Nature Human Behaviour (2021) doi:10.1038/s41562-021-01056-1.

40. Omer, S. B., Yildirim, I. & Forman, H. P. Herd immunity and implications for SARS-CoV-2 control. Jama 324, 2095–2096 (2020).

41. Aschwanden, C. Five reasons why COVID herd immunity is probably impossible. Nature 591, 520–522 (2021).

42. McNeil, D. G. How much herd immunity is enough? The New York Times (2021).

43. Mceachan, R., Conner, M., Taylor, N. & Lawton, R. Prospective prediction of health-related behaviours with the theory of planned behaviour: A meta-analysis. Health Psychology Review 5, 97 (2011).

44. Larson, H. J., Cooper, L. Z., Eskola, J., Katz, S. L. & Ratzan, S. Addressing the vaccine confidence gap. The Lancet 378, 526–535 (2011).

45. Hornsey, M. J., Finlayson, M., Chatwood, G. & Begeny, C. T. Donald trump and vaccination: The effect of political identity, conspiracist ideation and presidential tweets on vaccine hesitancy. Journal of Experimental Social Psychology 88, 103947 (2020).

46. Bokemper, S. E., Huber, G. A., Gerber, A. S., James, E. K. & Omer, S. B. Timing of COVID-19 vaccine approval and endorsement by public figures. Vaccine 39, 825–829 (2021).

47. Åslund, A. Responses to the COVID-19 crisis in russia, ukraine, and belarus. Eurasian Geography and Economics 61, 532–545 (2020).

48. Burki, T. K. The russian vaccine for COVID-19. The Lancet Respiratory Medicine 8, e85– e86 (2020).

49. Milkman, K. L., et al. A mega-study of text-based nudges encouraging patients to get vaccinated at an upcoming doctor’s appointment. (2021).

50. Milkman, K. L., et al. A mega-study of text-message nudges encouraging patients to get vaccinated at their pharmacy. (2021).

51. Levine, G., Salifu, A., Mohammed, I. & Fink, G. Mobile Nudges and Financial Incentives to Improve Coverage of Timely Neonatal Vaccination in Rural Areas (GEVaP trial): A 3-armed Cluster Randomized Controlled Trial in Northern Ghana. PLOS One (2021).

52. Gibson, D. G., et al. Mobile phone-delivered reminders and incentives to improve childhood immunisation coverage and timeliness in Kenya (M-SIMU): a cluster randomised controlled trial. The Lancet Global Health 5, e428–e438 (2017).

53. ID Insight. Impact of conditional cash transfers on routine childhood immunizations in North West Nigeria. https://files.givewell.org/files/DWDA%202009/NewIncentives/IDinsight_Impact_Evaluation_of_New_Incentives_Final_Report.pdf (2020).

54. Banerjee, A. V., Duflo, E., Glennerster, R. & Kothari, D. Improving immunisation coverage in rural India: clustered randomised controlled evaluation of immunisation campaigns with and without incentives. BMJ 340, (2010).

55. Brewer, N. T., Chapman, G. B., Rothman, A. J., Leask, J. & Kempe, A. Increasing vaccination: Putting psychological science into action. Psychological Science in the Public Interest 18, 149–207 (2017).

56. Betsch, C., Böhm, R. & Korn, L. Inviting free-riders or appealing to prosocial behavior? Game-theoretical reflections on communicating herd immunity in vaccine advocacy. Health Psychology 32, 978 (2013).

57. World Health Organization. Behavioural considerations for acceptance and uptake of COVID-19 vaccines: WHO technical advisory group on behavioural insights and sciences for health, meeting report, 15 October 2020. https://apps.who.int/iris/handle/10665/337335 (2020).

58. Katzman, J. G. & Katzman, J. W. Primary care clinicians as COVID-19 vaccine ambassadors. Journal of Primary Care & Community Health 12, 21501327211007026 (2021).

59. Alatas, V., Chandrasekhar, A. G., Mobius, M., Olken, B. A. & Paladines, C. When celebrities speak: A nationwide twitter experiment promoting vaccination in indonesia. National Bureau of Economic Research (2019).

60. Argote, P. et al. Messaging interventions that increase COVID-19 vaccine willingness in latin america. SSRN 3812023 (2021).

61. Beaman, L., BenYishay, A., Magruder, J. & Mobarak, A. M. Can network theory-based targeting increase technology adoption? National Bureau of Economic Research (2018).

62. Karing, A. Social signaling and childhood immunization: A field experiment in sierra leone. *University of California*, Berkeley Working Paper (2018).

63. World Bank. World Development Indicators: Life expectancy at birth, total (years) - Low & middle income, Low income countries [Data file]. https://data.worldbank.org/indicator/SP.DYN.LE00.IN?locations=XO-XM (2018).

64. World Bank. World Bank staff estimates based on age/sex distributions of United Nations Population Division’s World Population Prospects: 2019 Revision, Population ages 65 and above (% of total population) - Sub-Saharan Africa [Data file]. https://data.worldbank.org/indicator/SP.POP.65UP.TO.ZS?locations=ZG (2019).

65. Bank, W. World development report 2018: Learning to realize education’s promise. (The World Bank, 2017).

66. The Economist Intelligence Unit. Q1 global forecast 2021 Coronavirus vaccines: expect delays. https://www.eiu.com/n/campaigns/q1-global-forecast-2021/ (2021).

67. Centre for Disease Control and Prevention. Local Reactions, Systemic Reactions, Adverse Events, and Serious Adverse Events: Pfizer-BioNTech COVID-19 Vaccine. https://www.cdc.gov/vaccines/covid-19/info-by-product/pfizer/reactogenicity.html (2020).

68. CDC COVID-19 Response Team & Food and Drug Administration. Allergic Reactions Including Anaphylaxis After Receipt of the First Dose of Pfizer-BioNTech COVID-19 Vaccine — United States, December 14–23, 2020. https://www.cdc.gov/mmwr/volumes/70/wr/mm7002e1.htm#:~:text=Early%20safety%20monitoring%20of%20the,nonanaphylaxis%20allergic%20reactions%2C%20based%20on (2021).

69. Bowles, J., Larreguy, H. & Liu, S. Countering misinformation via WhatsApp: Preliminary evidence from the COVID-19 pandemic in zimbabwe. PloS one 15, e0240005 (2020).

